# General practice antibiotic prescriptions attributable to Respiratory Syncytial Virus by age and antibiotic class: An ecological analysis of the English population

**DOI:** 10.1101/2024.10.31.24316265

**Authors:** Lucy Miller, Thomas Beaney, Russel Hope, Mark Cunningham, Julie V. Robotham, Koen B. Pouwels, Cèire E. Costelloe

## Abstract

**Background:** Respiratory syncytial virus (RSV) may contribute to a substantial volume of antibiotic prescriptions in primary care. However, data on the type of antibiotics prescribed for such infections is only available for children <5 years in the UK. Understanding the contribution of RSV to antibiotic prescribing would facilitate predicting the impact of RSV preventative measures on antibiotic use and resistance.

**Objectives:** To estimate the proportion of antibiotic prescriptions in English general practice attributable to RSV by age and antibiotic class.

**Methods:** Generalised additive models examined associations between weekly counts of general practice antibiotic prescriptions and laboratory-confirmed respiratory infections from 2015 to 2018, adjusting for temperature, practice holidays and remaining seasonal confounders. We used general practice records from the Clinical Practice Research Datalink and microbiology tests for RSV, influenza, rhinovirus, adenovirus, parainfluenza, human Metapneumovirus, *Mycoplasma pneumoniae* and *Streptococcus pneumoniae* from England’s Second Generation Surveillance System.

**Results:** An estimated 2.1% of antibiotics were attributable to RSV, equating to an average of 640,000 prescriptions annually. Of these, adults ≥75 years contributed to the greatest volume, with an annual average of 149,078 (95% credible interval: 93,733-206,045). Infants 6-23 months had the highest average annual rate at 6,580 prescriptions per 100,000 individuals (95% credible interval: 4,522-8,651). Most RSV-attributable antibiotic prescriptions were penicillins, macrolides or tetracyclines. Adults ≥65 years had a wider range of antibiotic classes associated with RSV compared to younger age groups.

**Conclusions:** Interventions to reduce the burden of RSV, particularly in older adults, could complement current strategies to reduce antibiotic use in England.

## Introduction

Optimising antibiotic use by reducing unnecessary prescriptions and ensuring provision for those needing treatment are essential priorities to mitigate the significant threat antibiotic-resistant infections pose to healthcare. The UK’s 2024 National Action Plan aims to reduce total human antibiotic use by 5% by 2029 from a 2019 baseline.^1^ It has been suggested that respiratory syncytial virus (RSV) may generate a considerable number of primary care antibiotic prescriptions in the UK,^2–4^ most of which are anticipated to be unnecessary, given RSV presentations in primary care are typically self-limiting.^5,6^

Several RSV prophylactics, including vaccines and monoclonal antibodies (mABs) targeting infants, pregnant women, and older adults, have been licensed in the UK.^7^ These interventions may considerably reduce the burden of RSV and subsequent antibiotic use, which could impact resistant infections downstream.^7,8^ A secondary analysis of a trial indicated that a maternal vaccine could prevent 3.9 courses of antibiotics per 100 infants during their first year of life.^9^ However, vaccines for older adults in the UK may offer a greater impact, as this group has the highest rates of general practice (GP) antibiotic prescriptions^10^ and a considerable health burden from RSV.^3,11^

Understanding which population groups’ antibiotic prescribing is most likely affected by these programmes is important for informing their implementation and strategies to reduce antimicrobial resistance (AMR). Models which can predict the impact of programmes on antibiotic prescribing and subsequent resistant infections, incorporating the specific types of antibiotics likely to be reduced are needed, as it is well established that antibiotics vary in their selection for resistance.^12,13^ However, evidence of RSV-attributable antibiotic prescribing described by antibiotic type is only available for children <5 years in the UK.^4^

In this ecological study, we aimed to estimate the proportion of antibiotic prescriptions in English GPs attributable to RSV by age and antibiotic class.

## Methods

### Ethics

The study protocol was approved by the Clinical Practice Research Datalink (CPRD) Independent Scientific Advisory Committee (protocol 20_000283) and the Imperial College Research Ethics Committee (reference number 21IC6607).

### Study period and data

We used ecological regression analyses to estimate weekly antibiotic prescriptions attributable to RSV from 29 December 2014 to 30 December 2018, using weekly counts of laboratory-confirmed respiratory infections and average weekly temperatures as explanatory covariates. The study period was selected to avoid the impact of the COVID-19 pandemic on antibiotic prescribing and respiratory infections.^14–16^

Data on antibiotic prescriptions were obtained from CPRD Aurum, a dataset containing anonymised electronic health records from contributing GPs in England. This dataset represents ∼23% of the English population^17,18^ and broadly reflects national demographics regarding age, sex, and deprivation.^19^ Records of research-acceptable patients registered between 1 January 2015 and 1 January 2020 and linked to hospital records from the Hospital Episode Statistics (HES) database and practice-level Index of Multiple Deprivation (IMD) were extracted.^20^ CPRD determines research acceptability based on data reliability, including date of birth, practice registration date and transfer out date.^21^ The study period began three days before the extraction period, which excluded patients registered only during those days, with minimal expected data loss. Linkage to HES and IMD, required for analyses outlined in the CPRD protocol, resulted in the exclusion of approximately 2.5% of research-acceptable patients.

Antibiotic prescriptions were identified irrespective of the presenting condition, as up to a third could have missing or non-specific diagnostic codes in English primary care.^22^ Antibiotics included any systemic antibacterial (J01) listed in the Anatomical Therapeutic Chemical (ATC) classification system^23^ and Chapter 5.1 of the British National Formulary (BNF),^24^ excluding those used for leprosy, tuberculosis, and topical applications, except for those recommended for ear infections.^6^ The primary outcomes included respiratory antibiotics primarily used for respiratory tract infections (RTIs),^6,22^ namely amoxicillin, phenoxymethylpenicillin, clarithromycin, erythromycin and doxycycline, along with antibiotic class groups defined by the BNF Chapter 5.1 subsections.^24^ Secondary outcomes included total antibiotic prescriptions and respiratory antibiotics of potential importance for resistance to assess RSV’s contribution to overall antibiotic use across different age groups and the resistance propensity of these prescriptions. Outcomes are defined in Section 1 and Table S.1 of Supplementary data. Nitrofurantoin prescriptions typically used only for UTIs,^22,25^ were analysed as a negative control to identify any unaccounted residual confounding. All prescriptions were aggregated by calendar week and stratified by age: 0-5 months, 6-23 months, 2-4 years, 5-14 years, 15-44 years, 45-64 years, 65-74 years, and ≥75 years.

Positive laboratory tests of respiratory pathogens in the English population, including RSV, influenza, rhinovirus, adenovirus, parainfluenza, human Metapneumovirus (hMPV), *Mycoplasma pneumoniae* and *Streptococcus pneumoniae,* were extracted from the UK Health Security Agency’s Second Generation Surveillance System (SGSS). The SGSS collects routine infectious disease test results from around 98% of hospital laboratories in England.^26,27^ Tests are voluntarily submitted by healthcare professionals, with clinically significant infections likely captured and most culture requests originating from hospitals. We included all respiratory samples for viruses and respiratory and invasive samples for *M. pneumoniae*. Only invasive samples were included for *S. pneumoniae* due to inconsistent reporting of respiratory samples.^27^ Tests from the same patient for the same pathogen were grouped if reported within two weeks (six weeks for influenza) to prevent over-reporting.^27^ Tests were then aggregated by calendar week and stratified by broad age groups (0-4 years, 5-64 years, and ≥65 years) to adjust for age-specific differences in the seasonality of infections.^28^

Daily average temperatures for England were obtained from the Hadley Centre Central England Temperature dataset (HadCET)^29^ and averaged by calendar week.

### Statistical analysis

Separate models were developed for each outcome by age, associating outcomes with the corresponding counts of laboratory-confirmed infections in broad age groups and average temperatures. We explored the seasonality of antibiotic prescriptions and laboratory-confirmed infections and used correlation matrices to examine the collinearity between pathogens and temperature.

We fitted generalised additive models (GAMs) with a negative binomial distribution and an identity link. The negative binomial distribution accounted for overdispersion in outcome counts, while the identity link ensured each laboratory-confirmed infection contributed additively to these counts. GAMs allowed for non-linear covariate relationships using splines, fitting data-derived trends using restricted maximum likelihood estimation (REML),^30^ providing flexible adjustment for unmeasured seasonal confounding. Splines were expanded to penalise covariates with no relationship for variable selection (double penalty approach), enabling penalisation for deviations from a straight line and straight-line components to be shrunk to zero.^30^ To adjust for an increasing CPRD population, all covariates, including the intercept, were multiplied by the average mid-year CPRD population of the relevant age group and calendar week, effectively applying an offset (Table S.2 and Equation S.1). This method preserves the scale in identity link models, enabling straightforward interpretation of the results as absolute changes in outcome counts.

Positive tests, likely from hospitalised patients, were assumed to reflect community incidence, as both are expected to occur within approximately a week of each other. For instance, paediatric RSV hospitalisations are reported to occur 3–4 days after symptom onset,^31^ and the average duration of RTI symptoms is estimated to be around 3–14 days.^32,33^ Weekly lags were applied to align the seasonality of confirmed RSV infections across broad age groups with age groups used for outcomes. This alignment was evaluated using autocorrelation function (ACF) plots. The Akaike information criterion (AIC) tested the inclusion of 3-week moving averages of pathogen counts, linear and non-linear outcome trends, and practice holiday indicator variables. Pathogen counts and temperatures were fitted with penalised splines using REML with a double penalty to explore non-linear trends and conduct variable selection.

GAMs were performed using the “mgcv” package in R version 4.2.1.^34^ Model fit was assessed by comparing posterior simulations with observed data. Posterior simulations were generated using the “postSim” function from the mgcViz package,^35^ ensuring they were drawn from a multivariate normal distribution defined by the model’s variance-covariance matrix, incorporating uncertainties and correlations among covariates.^35,36^ Model stationary was assessed using ACF, residual, and quantile-quantile plots.

RSV-attributable prescriptions were estimated by subtracting posterior simulations of weekly outcome counts from a model with RSV counts set to zero, from those of a model using the observed RSV counts. Simulations were repeated 1000 times, taking the 2.5% and 97.5% percentiles to estimate 95% credible intervals (CrI). Prescribing rates were estimated using age-specific mid-year CPRD study population estimates and scaled to the national level with age-specific ONS mid-year English population estimates between 2015 and 2018 (Table S.2).^18^ Average age-specific counts of RSV-attributable antibiotic classes were used to estimate age-specific class proportions of RSV-attributable antibiotic prescriptions. Only class counts with an estimated 2.5% percentile above zero were considered in this estimation, excluding UTI antibiotic prescriptions.

### Sensitivity analysis

All respiratory pathogens included in the GAMs have evidence of potential co-infections with RSV,^28,37–40^ which could lead to underestimating RSV’s contribution if RSV increases hosts’ susceptibility to other pathogens. To address this, we evaluated the impact of excluding pathogens highly correlated with RSV (>0.7) on estimated age-specific RSV-attributable proportions. Furthermore, we assessed the impact of removing all respiratory pathogens except influenza and RSV on age-specific RSV-attributable proportions, as previous studies of RSV-attributable antibiotic prescriptions in the UK have only controlled for influenza.^2,3^

## Results

Over the 4-year study period, 27,969,054 antibiotic prescriptions from 17,505,438 CPRD-registered patients were analysed, with 192,938 laboratory-confirmed respiratory infections (36,180 = RSV) and average weekly temperatures of median 10.3°C (IQR 6.6-14.6) (Tables S.3-4). Antibiotic and respiratory antibiotic prescription rates per year decreased from 584 and 287 per 1,000 individuals in 2015 to 501 and 232 per 1,000 individuals in 2018 (Table S.3). Respiratory antibiotics comprised approximately half of all antibiotic prescriptions (Table 1), with penicillins most frequently prescribed for all ages (Figure 1 and Table S.4). Infants 6-23 months had the highest rate of respiratory antibiotic prescriptions, while adults ≥75 years had the highest rate of antibiotic prescriptions (Table 1). Antibiotic and respiratory antibiotic prescriptions demonstrated winter seasonality, mainly driven by penicillins (Figure 1).

**Table 1:** Average annual antibiotic prescriptions and prescriptions per 100,000 in CPRD by age from 29 December 2014 to 30 December 2018. % = The age-specific proportion of outcome counts out of total antibiotic prescriptions. Respiratory antibiotic prescriptions included amoxicillin, phenoxymethylpenicillin, clarithromycin, erythromycin and doxycycline. Respiratory antibiotic prescriptions potentially important for resistance included amoxicillin, co-amoxiclav, phenoxymethylpenicillin, flucloxacillin, cefalexin, doxycycline, gentamicin, erythromycin, clarithromycin, azithromycin, levofloxacin, ciprofloxacin and co-trimoxazole (Table S.1).

| Age | Antibiotic prescriptions |  | Respiratory antibiotic prescriptions |  |  | Respiratory antibiotic prescriptions potentially important for resistance |  |  | Nitrofurantoin prescriptions (negative control) |  |  |
| --- | --- | --- | --- | --- | --- | --- | --- | --- | --- | --- | --- |
|  | Number | Rate per 100,000 | Number | Rate per 100,000 | % | Number | Rate per 100,000 | % | Number | Rate per 100,000 | % |
| 0-5 months | 11,712 | 30,462 | 7,250 | 18,849 | 62 | 10,202 | 26,528 | 87 | 29 | 75 | <1 |
| 6-23 months | 190,175 | 79,088 | 156,692 | 65,164 | 82 | 181,445 | 75,458 | 95 | 422 | 176 | <1 |
| 2-4 years | 283,890 | 59,847 | 216,771 | 45,696 | 76 | 258,962 | 54,591 | 91 | 1,006 | 212 | <1 |
| 5-14 years | 462,970 | 31,488 | 286,549 | 19,506 | 62 | 384,153 | 26,410 | 83 | 5,180 | 350 | 1 |
| 15-44 years | 1,940,258 | 37,319 | 860,118 | 16,556 | 44 | 1,287,830 | 24,781 | 66 | 167,271 | 3,191 | 9 |
| 45-64 years | 1,711,041 | 52,381 | 810,115 | 24,816 | 47 | 1,196,182 | 36,633 | 70 | 158,714 | 4,832 | 9 |
| 65-74 years | 1,022,016 | 87,552 | 473,456 | 40,577 | 46 | 715,019 | 61,269 | 70 | 110,386 | 9,417 | 11 |
| ≥75 years | 1,370,201 | 136,942 | 546,854 | 54,669 | 40 | 890,511 | 89,015 | 65 | 187,095 | 18,627 | 14 |
| Total in CPRD | 6,992,263 |  | 3,357,805 |  | 48 | 4,924,304 |  | 70 | 630,103 |  | 9 |

**Figure 1:**
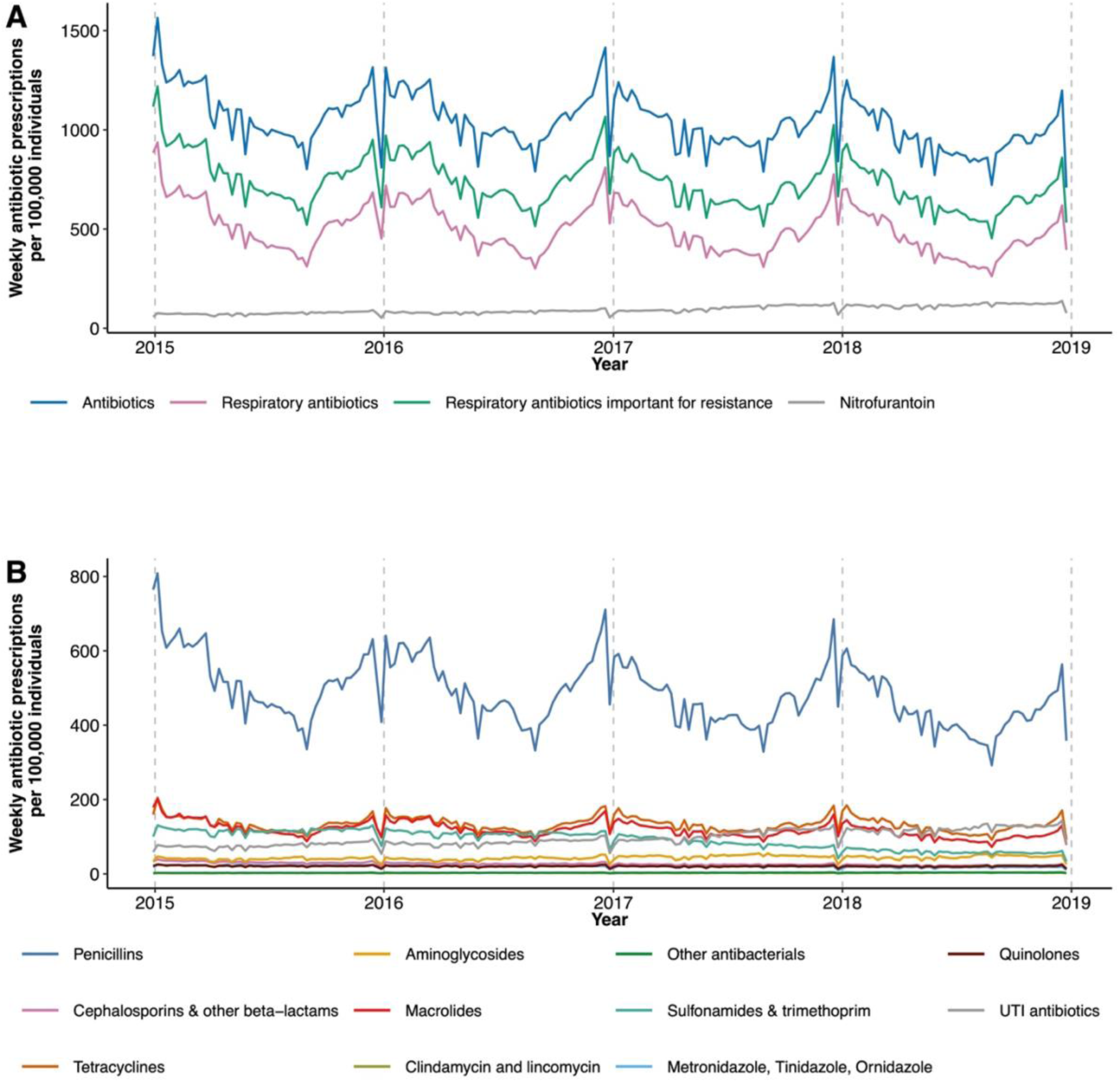
Weekly antibiotic prescriptions per 100,000 individuals in CPRD from 29 December 2014 to 30 December 2018. A = Antibiotic prescription outcomes, B= Classes of antibiotic prescriptions.

Figures 2 and 3 demonstrate the seasonality of RSV and respiratory pathogens across three broad age groups: 0-4 years, 5-64 years, and ≥65 years. Around 74% of laboratory-confirmed RSV infections came from children <5 years, with sharp winter peaks observed for all ages (Figure 2). Peaks occurred later with increasing age, from late November for children <5 years to early January for adults ≥65 years (Figure 2). RSV dominated laboratory-confirmed respiratory infections from children <5 years during early winter, while influenza constituted most infections from individuals ≥5 years during late winter (Figure 3). Across most age groups used for antibiotic prescriptions, peaks of RSV infections aligned with those of their broader age groups. RSV infections for 5–14 years peaked one week earlier than in 5-64 years (Figure S.1).

**Figure 2:**
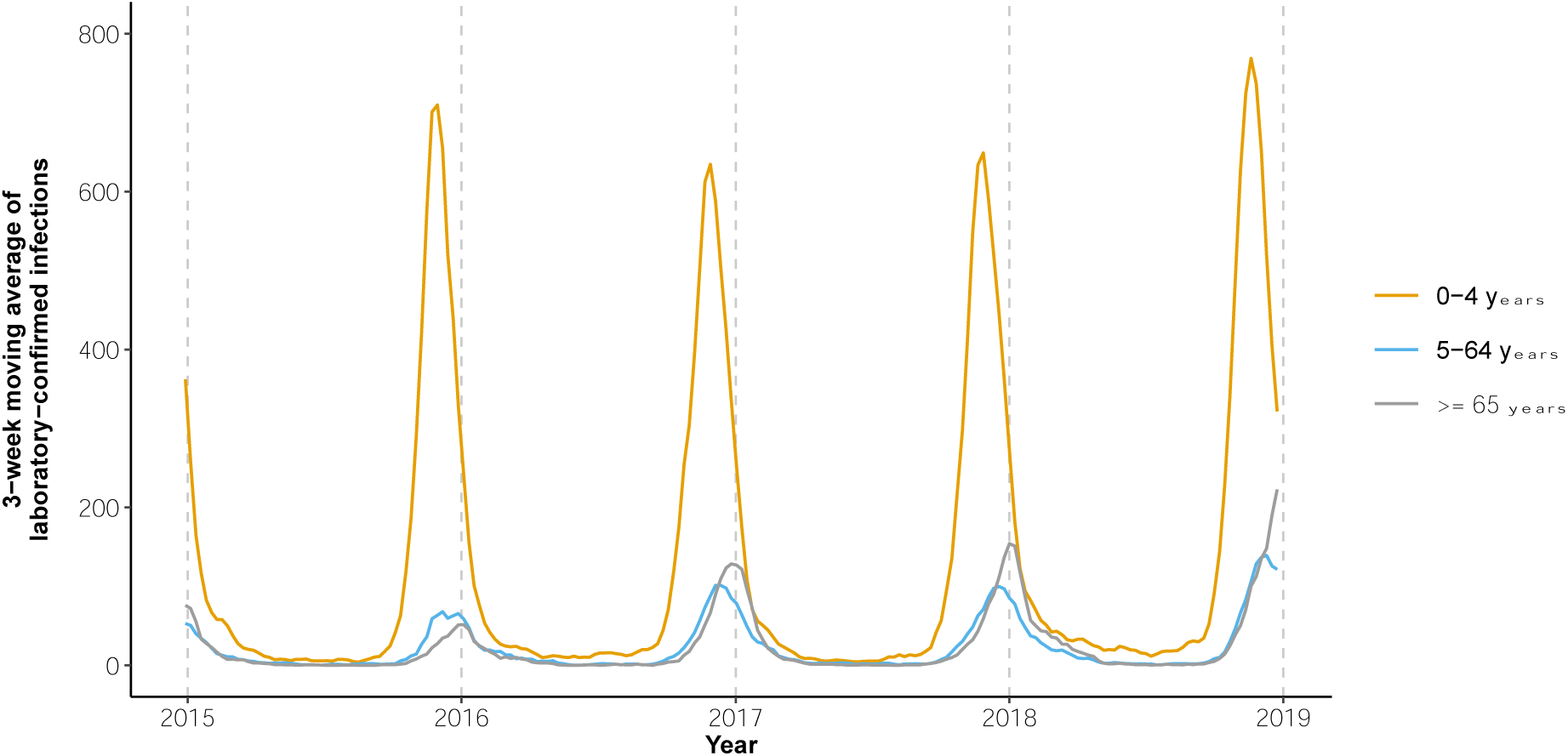
Three-week moving averages of laboratory-confirmed RSV infections in England recorded in SGSS from 29 December 2014 to 30 December 2018 stratified by age.

**Figure 3:**
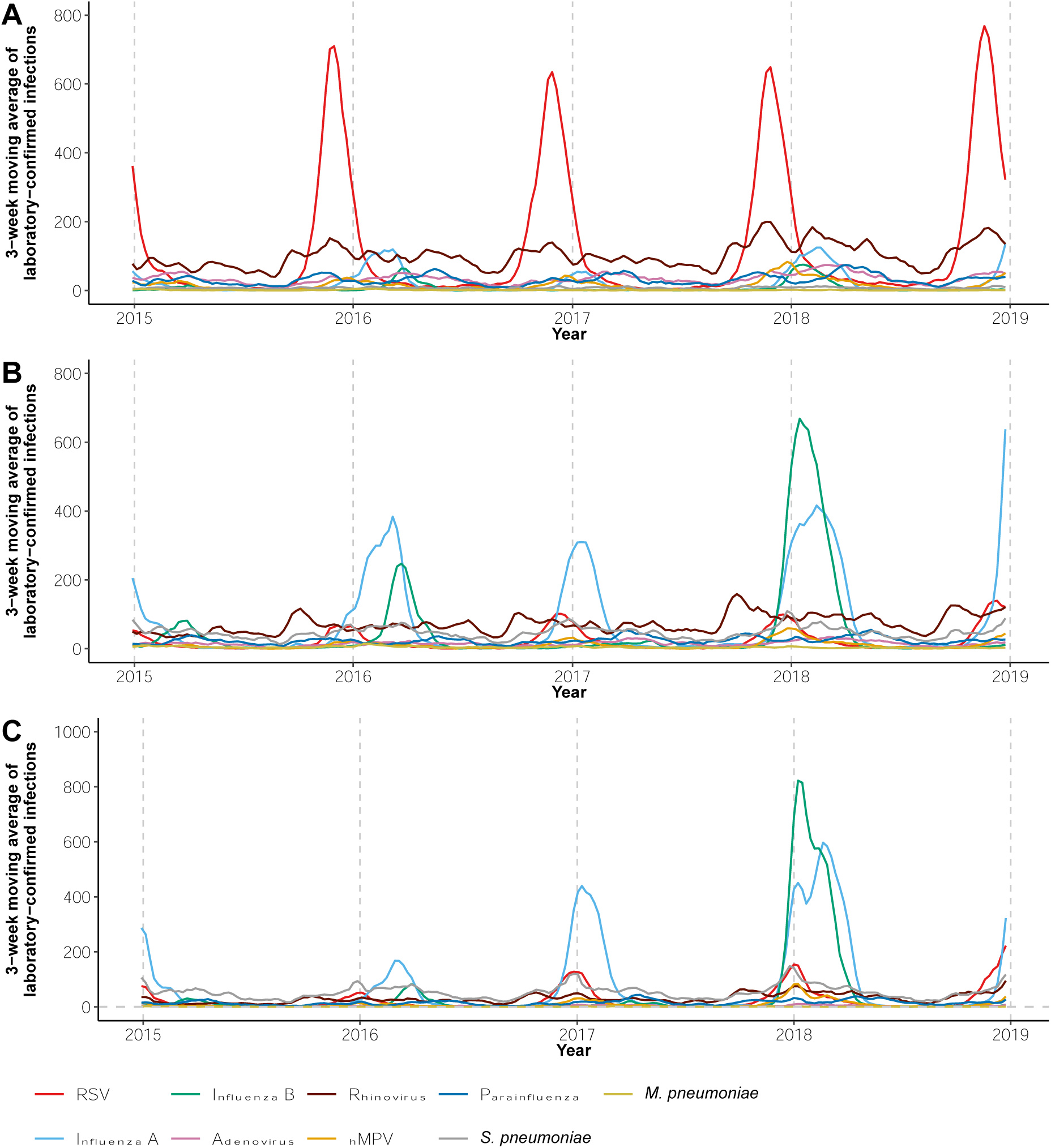
Three-week moving averages of laboratory-confirmed respiratory infections in England recorded in SGSS from 29 December 2014 to 30 December 2018 for individuals aged 0-4 years (A), 5-64 years (B) and ≥65 years (C).

Weekly respiratory infections of broad age groups demonstrated negative correlation with average temperatures in England (Figure S.2). Most pathogens had low to moderate correlation with RSV across all age groups (Figure S.2). High collinearity (>0.7) with RSV was observed for hMPV and *S. pneumoniae* in adults ≥65 years and for hMPV in individuals 5-64 years.

### Model fitting

The best-fitting model for all ages included a moving average to smooth irregularities in pathogen counts, all practice holiday indicator variables to adjust for outliers in outcome counts, and a spline to account for unmeasured seasonal confounding (Figure S.3). Models for adults ≥65 and children 5-14 years indicated potential non-linear relationships between pathogens (including influenza, RSV, hMPV, and *S. pneumoniae*) and outcomes, influenced by a few outliers in pathogen counts with high uncertainty (Figure S.4). To address this, weekly counts of respiratory infections above the 97.5th percentile were truncated to the 97.5% value for age-specific models (Figure S.4). The final set of time-varying covariates varied between age-specific models (Figure S.5), with RSV, influenza, rhinovirus, and *S. pneumoniae* frequently included in models of respiratory antibiotic prescriptions. After truncation, the trend of remaining time-varying covariates was approximately linear (Figure S.4); therefore, splines were removed to reduce unnecessary uncertainty.

Model residuals demonstrated reasonable normality with minimal heteroskedasticity or autocorrelation (Figure S.6). Possible autocorrelation was noted in models for 0-5 months, 2-4 years and 5-14 years. However, attempts to adjust for this using Gaussian process splines^41^ suggested insufficient remaining residual trend for autocorrelation (Figure S.7). Most posterior simulations of age-specific models of respiratory antibiotic prescriptions closely matched observed data (Figures S.8-10), except for 0-5 months, which reflected an average of 2-4 weekly fluctuations previously noted in Scottish children.^4^ The 5-14 years model struggled to simulate a no-RSV scenario in 2018 (Figure S.11). To address this, the model was adjusted by excluding 2018 data and decomposing seasonal trends into two splines to better capture long-term and repeating seasonal patterns.^42^

### RSV-attributable antibiotic prescribing

An estimated 2.1% of all antibiotic prescriptions and 4.3% of respiratory antibiotic prescriptions were attributable to RSV infections across all ages, amounting to an annual average of 639,908 GP prescriptions in England. Infants 6-23 months had the highest rates of RSV-attributable prescriptions, with an annual average of 6,580 prescriptions per 100,000 individuals (95% CrI: 4,522-8,651) (Table 3). Adults ≥75 years had the highest annual volume of RSV-attributable prescriptions at 149,078 (95% CrI: 93,733-206,045). Secondary outcomes demonstrated a greater pool of antibiotic prescriptions, beyond those typically used for RTIs, associated with RSV infections in adults ≥45 years, with most attributable prescriptions across all ages likely important for resistance selection and development (Table S.5).

**Table 2:** Average annual RSV-attributable GP respiratory antibiotic prescriptions in England from 29 December 2014 to 30 December 2018 stratified by age. % = age-specific proportion. CrI = credible interval, m = months, y = years. *= Prescriptions are estimated assuming English mid-year populations are equally distributed by month of age as ONS population estimates are only provided by year of age. # = Average annual RSV-attributable prescriptions for the 5-14 age group were estimated from 2015 to 2017 and assumed to apply to 2018, as 2018 data was excluded for this group (see model fitting).

| **Average annual RSV-attributable GP respiratory antibiotic prescriptions in England** |  |  |  |  |
| --- | --- | --- | --- | --- |
| **Age** | **Prescriptions (95% CrI)** | **%** | **Attributable proportion % (95% CrI)** | **Rate per 100,000 (95% CrI)** |
| **0-5 m** | 6,880* (3,211-10,167) | 1.1 | 11 (5-17) | 2,100 (984-3,100) |
| **6-23 m** | 65,535* (45,035-86,144) | 10.2 | 10 (7-13) | 6,580 (4,522-8,651) |
| **2-4 y** | 72,500 (29,222-119,821) | 11.3 | 8 (3-13) | 3,497 (1,405-5,782) |
| **5-14 y#** | 55,860 (-41,759-156,160) | 8.7 | 4 (-3-12) | 857 (-637-2,397) |
| **15-44 y** | 95,554 (6,792-185,445) | 14.9 | 3 (0-5) | 447 (31-868) |
| **45-64 y** | 86,608 (-16,608-191,877) | 13.5 | 2 (0-6) | 612 (-117-1,357) |
| **65-74 y** | 107,893 (60,407-158,507) | 16.9 | 5 (3-7) | 1,972 (1,104-2,901) |
| **≥75 y** | 149,078 (93,733-206,045) | 23.3 | 6 (4-8) | 3,279 (2,050-4,532) |

Adults ≥65 years had a broader spectrum of antibiotic classes associated with RSV than younger age groups (Table 3). Among children <5 years, penicillins and macrolides had the highest proportion attributable to RSV, with 10% (95% CrI: 6-14) and 8% (95% CrI: 4-11), respectively, for infants 6-23 months. For adults ≥65 years, tetracyclines had the highest proportion attributable to RSV, with 6% (95% CrI: 3-8) for ≥75 years. Across all ages, penicillins accounted for the most RSV-attributable prescriptions, followed by macrolides and tetracyclines (Figure S.12).

**Table 3:** Age-specific RSV attributable GP antibiotic prescriptions by class described by RSV attributable proportion. m = months, y = years, CrI = credible interval, PEN = Penicillin’s, CEPH+ = Cephalosporins & other beta lactams, TET = Tetracyclines, AMINO = Aminoglycosides, MAC = Macrolides, CLI+ = Clindamycin & Lincomycin, OTHER = Other antibacterials, SULF+ = Sulfonamides & Trimethoprim, MTZ+ = Metronidazole, Tinidazole & Ornidazole, QUIN = Quinolones, UTI = Urinary tract infection antibiotics, - = The model was not run because age-specific counts of antibiotic classes were <1,000 during the study period or demonstrated no relationship with confirmed RSV infections.

| **Age-specific RSV attributable proportion % of GP antibiotic prescriptions by class (95% CrI)** |  |  |  |  |  |  |  |  |
| --- | --- | --- | --- | --- | --- | --- | --- | --- |
| **Class** | **0-5 m** | **6-23 m** | **2-4 y** | **5-14 y** | **15-44 y** | **45-64 y** | **65-74 y** | **≥ 75 y** |
| **PEN** | 9 (4-14) | 10 (6-14) | 7 (2-11) | 3 (-4-11) | - | 3 (1-5) | 3 (1-6) | 4 (2-6) |
| **CEPH+** | - | 3 (-2-7) | 4 (0-8) | - | - | - | 2 (0-5) | 2 (0-4) |
| **TET** | - | - | - | - | - | 3 (0-5) | 5 (2-7) | 6 (3-8) |
| **AMINO** | - | -4 (-14-5) | - | - | 2 (-1-6) | 2 (-1-5) | 4 (1-6) | 4 (1-7) |
| **MAC** | 2 (-8-11) | 8 (4-11) | 7 (3-10) | 3 (-3-10) | - | 2 (0-4) | 4 (2-7) | 5 (2-7) |
| **CLI+** | - | - | - | - | - | - | - | - |
| **OTHER** | - | - | - | - | - | 1 (-2-4) | 4 (0-7) | - |
| **SULF+** | 1 (-5-8) | - | 1 (-2-4) | - | - | - | 2 (0-4) | 2 (0-4) |
| **MTZ+** | - | - | - | - | 1 (-2-3) | 1 (-1-3) | 2 (-1-4) | 1 (-1-4) |
| **QUIN** | - | - | - | - | - | - | - | 3 (0-5) |
| **UTI** | - | - | - | - | 4 (0-7) | 3 (0-5) | 2 (0-5) | 2 (0-5) |

### Negative control and sensitivity analysis

The negative control analysis with nitrofurantoin prescriptions, exclusively for UTIs, as the outcome demonstrated a potential association with RSV infections in individuals 15-44 years and ≥75 years (Table S.6 and Figure S.13).

Removing *S. pneumoniae* from models for adults ≥65 years and hMPV from models for 5-14 and 45-64 years, which were highly correlated with RSV infections, increased the estimated RSV contribution by one percentage point for adults ≥45 years (Table S.7). Removing all pathogens apart from influenza and RSV increased the estimated RSV contribution by 1-14 percentage points for most ages, with the largest increase in children <5 years (Table S.7).

## Discussion

### Principal findings

Our analyses estimated that 2.1% of antibiotic prescriptions in English GPs were attributable to RSV infections. Prescribing to adults ≥75 years contributed to the greatest degree, despite infants between 6-23 months having the highest estimated rate of RSV-attributable prescribing. This was driven by the greater population size and high antibiotic prescribing rates of older adults (Table 1). The antibiotic classes frequently attributed to RSV infections were those recommended for RTIs, e.g., penicillins, macrolides and tetracyclines.^6^ However, our study suggested that a broader spectrum of antibiotic classes was associated with RSV infections in older adults, potentially due to the increased challenges of diagnosing infections in this age group.^43^

### Strengths and weaknesses

To our knowledge, this study provides the first estimate of RSV-attributable primary care antibiotic prescriptions by antibiotic class for individuals ≥5 years in the UK, using nationally representative GP records and laboratory-confirmed respiratory infections. We controlled for a wide range of respiratory pathogens that could drive RTI antibiotic prescribing and stratified counts by age to reflect age-specific differences in pathogen seasonality (Figure 2-3). This was important given the higher correlation between respiratory infections in older adults (Figure S.2). A key strength of our approach lies in using GAMs, which provided robust variable selection and fitting. GAMs effectively penalised variables unrelated to antibiotic prescribing while considering other relationships in a single step, minimising bias typically introduced by popular stepwise regression techniques.^30^ Additionally, GAMs allowed for more flexible adjustment of unmeasured seasonal confounding by fitting data-derived trends ^30^ instead of assuming fixed cyclic patterns that could introduce bias.

We identified three previous studies estimating RSV-attributable antibiotic prescribing in the UK.^2–4^ Taylor *et al*.^2^ and Fleming *et al*.^3^ using CPRD data from 1995-2009 and only controlling for influenza, reported higher proportions of RSV-attributable prescribing compared to our analysis. They estimated that 14.6% of respiratory antibiotic prescriptions in infants 6-23 months were attributable to RSV, compared to 11% in our study. In adults 64-75 years and ≥75 years, they found 6% and 6.3% of respiratory antibiotic prescriptions attributable to RSV compared to our estimates of 5% and 6%. Our more conservative estimates likely reflect adjustments for additional respiratory pathogens in younger age groups, where controlling for pathogens beyond influenza was suggested to decrease RSV-attributable antibiotic prescriptions by up to 14 percentage points in children <5 years (Table S.7). Additionally, GP antibiotic prescriptions significantly declined between study periods.^44^

Fitzpatrick *et al.* ^4^ analysed data for children <5 years in Scotland from 2009-2017, associating laboratory tests of multiple respiratory pathogens with all community antibiotic prescriptions, including those from additional providers like dentists. They reported slightly lower proportions of RSV-attributable antibiotic prescriptions for children <5 years (5.8%) compared to our secondary analysis of GP-only prescriptions (6-8%, Table S.5). This discrepancy may stem from Fitzpatrick *et al.* ^4^ using a larger denominator of all community antibiotic prescriptions, potentially diluting the attributable proportion. Scaling our results to average community antibiotic prescribing for the study period gives a comparable 5.2-6.9% for children <5 years.^44^ They also estimated comparable RSV-attributable proportions for penicillin (amoxicillin) and macrolide prescriptions in this age group (8.1% and 7.7% vs 7-10% in our study). Our study is the first to estimate age-specific RSV-attributable proportions for antibiotic classes beyond those typically prescribed for RTIs and across all ages.

Our study’s annual rates of antibiotic and respiratory antibiotic prescriptions in GPs were slightly lower than previously reported for 2015-2018. The English Surveillance Programme for Antimicrobial Utilisation and Resistance (ESPAUR) report estimated 602 to 532 per 1,000 individuals in English GPs between 2015 and 2018, compared to our rates of 584 to 501 per 1,000 individuals.^44^ Similarly, national analyses of respiratory antibiotic prescriptions in the community, irrespective of prior diagnosis, estimated quarterly rates of 65-100 per 1,000 individuals,^16^ while our average quarterly rates were ∼58-72 per 1,000 individuals. These lower rates likely reflect our more conservative outcome definitions, which excluded topical antibiotics (except for ear infections) from the overall count and certain respiratory antibiotics, such as co-amoxiclav, commonly used for other conditions (Section 1 Supplementary data).^22^ Co-amoxiclav had quarterly community dispensing rates of ∼6-8.25 per 1,000 individuals during this period.^45^

Common limitations of studies utilising laboratory data of respiratory pathogens include underreporting and most tests likely being from hospital cases, which may not represent the study population. Biases may arise from changes in testing practices, healthcare-seeking behaviours, or discrepancies between our age groups for surveillance and prescription data. For example, the 5-14 years model struggled with 2018 data (Figure S.11), likely due to a significant increase in influenza samples in 5-64 years, more representative of adults (Figure 3).^28^

The study could not control for possible RSV co-infections, potentially underestimating RSV’s contribution. Sensitivity analysis suggested that including hMPV or *S. pneumoniae,* both highly correlated with RSV in adults ≥45 years, may have underestimated RSV-attributable antibiotic prescriptions by one percentage point in this age group. Co-infections with *S. pneumoniae* and hMPV are linked to increased disease severity,^38,46^ and RSV may enhance the susceptibility and virulence of *S. pneumoniae*,^47^ thus results for older adults may be conservative. However, co-infection frequency in the general population remains poorly understood, as most evidence is based on co-detections from symptomatic samples, which are prone to bias.^48,49^ Country-specific, community-based studies that collect respiratory pathogen samples regardless of clinical status,^48^ accounting for factors like viral load (to indicate infection),^49^ age, and comorbidities, are needed to understand infection and co-infection incidence better.

Finally, unmeasured confounding may lead to overestimated model predictions. The negative control demonstrated an association between RSV infections and nitrofurantoin prescriptions for individuals 15-44 and ≥75 years. In 15-44 years, this association may indicate unaccounted-for confounding due to overlapping seasonality of RTIs and UTIs at the start of university periods,^50^ suggesting a potential overestimation of the RSV contribution in this group. However, in adults ≥75 years, winter peaks in nitrofurantoin prescriptions do not likely match UTI activity.^50^ Instead, these peaks may be due to older adults with RTIs being treated for UTIs, reflecting non-specific symptoms and diagnostic challenges in identifying a source of infection,^51,52^ highlighting the difficulties of antibiotic stewardship in this group.^43,51^

### Implications and conclusions

Our study suggests that interventions like vaccines or mABs to reduce the burden of RSV infections in England could complement national efforts to reduce antibiotic use.^1^ The largest potential reductions are in older adults, an age group for whom antibiotic stewardship is challenging.^43,51^

The impact of RSV prescribing reductions on AMR remains unclear. Most RSV-attributable prescriptions were “Access” antibiotics, such as amoxicillin and doxycycline, recommended by the WHO as first and second-line treatments for common infections and considered to have lower resistance potential.^53^ However, extensively used antibiotics like amoxicillin may affect commensal pathogens systematically,^12^ with evidence of potentially promoting the co-selection of resistant UTIs in the community.^13,54^ This study was unable to explore secondary care antibiotic prescriptions, which, though smaller in volume, typically involve broader spectrum antibiotics with higher resistance potential^10^ and are used in patients at greater risk of severe resistant infections.

Despite these limitations, our findings provide a prerequisite for exploring the onward impacts of reduced RSV-related prescribing on AMR and can inform future modelling.

## Supporting information

Section 1, Figures S.1 to S.13, Equation S.1 and Tables S.1 to S.6 are available as supplementary data.

## Acknowledgements

Imperial College London is grateful for support from the National Institute for Health Research (NIHR) Imperial Biomedical Research Centre and the NW London NIHR Applied Research Collaboration. Data secure storage and management was provided by the Big Data and Analytical Unit (BDAU) at the Institute of Global Health Innovation. The authors are thankful to Dr Shirin Aliabadi and Dr Monsey McLeod for providing feedback on antibiotic therapy codes.

## Transparency declaration

### Author contributions

C.E.C, K.B.P and J.V.R conceptualised and supervised the project; they supported study design, analyses and interpretation of results. L.M conceptualised the project, designed the study, acquired the data, performed analysis, and wrote the original article. T.B supported study design and interpretation of results. R.H and M.C substantially contributed to the acquisition of data. All authors contributed to revising the original manuscript and approved the final version.

### Declaration of Generative AI and AI-assisted technologies in the writing process

During the preparation of this work the author used ChatGPT 4o to improve readability and language of the article. After using this tool, the author reviewed and edited the content as needed and takes full responsibility for the content of the publication.

### Data availability

This study is based in part on data from the Clinical Practice Research Datalink obtained under licence from the UK Medicines and Healthcare products Regulatory Agency. The data is provided by patients and collected by the NHS as part of their care and support. Data from the Second Generation Surveillance System was provided by UKHSA under licence. Both datasets are not publicly available. However, an application for CPRD data can be made to the Independent Scientific Advisory Committee and SGSS data can be requested via the office of data release at UKHSA.

### Disclaimer

The interpretation and conclusions contained in this study are those of the authors alone, and not necessarily those of the Medical Research Foundation, NIHR, Department of Health and Social Care, UKHSA or CPRD.

