## Supplementary material for "General practice antibiotic prescriptions attributable to Respiratory Syncytial Virus by age and antibiotic class: An ecological analysis of the English population": Section 1, Figures S.1 to S.13, Equation S.1 and Tables S.1 to S.6 are available as supplementary data.

**Section 1:** Definition of outcomes

Classes of antibiotics are defined by the British National Formulary (BNF) Chapter 5.1 subsections^1^ including;

- Penicillins
- Cephalosporins and other beta-lactams
- Tetracyclines
- Aminoglycosides
- Macrolides
- Clindamycin and Lincomycin
- Other antibacterials
- Sulfonamides and Trimethoprim
- Metronidazole, Tinidazole and Ornidazole
- Quinolones
- UTI antibiotics

Respiratory antibiotic prescriptions are specified as antibiotics that are most frequently used to treat respiratory tract infections (RTIs) in primary care with only a small overlap of use for other non-respiratory conditions,^2–8^ namely: amoxicillin, phenoxymethylpenicillin, clarithromycin, erythromycin and doxycycline. Respiratory antibiotics potentially important for resistance (Table S.1) were defined as antibiotics that are recommended in National Institute for Health and Care Excellence (NICE) treatment guidance for RTIs and ear infections,^2–8^ and have either:

i) Evidence of clinically relevant resistance in bacteria of high carriage prevalence in the UK, according to recent ESPAUR reports.
ii) Evidence of clinically relevant resistance as described in (i), where most of the bacteria’s exposure to the antibiotic is potentially from bystander exposure.^9^
iii) Evidence of driving clinically relevant resistance (i) to other antibiotics via co-selection.
iv) Evidence of a high volume of use in UK primary care, therefore a potential candidate of co-selection for organisms of high carriage prevalence in the UK.

Nitrofurantoin prescriptions typically only used for treating urinary tract infections (UTIs)^2,10^ were used as a negative to explore unaccounted-for residual confounding. A practising pharmacist and clinical microbiologist reviewed finalised lists of respiratory antibiotics and respiratory antibiotics potentially important for resistance. A practicing general practitioner reviewed finalised lists of antibiotic outcomes.

**Table S.1:** Finalised list of respiratory antibiotics potentially important for resistance in the UK

| **Respiratory antibiotics** | **Clinically relevant resistance in the UK** | | | | **Co-selection of resistance** | | **High volume** | **Reference** |
| --- | --- | --- | --- | --- | --- | --- | --- | --- |
|  | ***E. coli*** | ***K. pneumoniae*** | ***S. pneumoniae*** | ***S. aureus*** | ***E. coli*** | ***S. aureus*** |  |  |
| **Penicillins** | | | | | | | |  |
| Co-amoxiclav | **x** | **x** | **x** |  |  |  |  | ^11,12^ |
| Amoxicillin |  |  | **x** |  | **x** |  |  | ^2,11–14^ |
| Phenoxymethylpenicillin |  |  |  |  |  |  | **x** | ^2,11,12^ |
| Flucloxacillin |  |  |  |  |  |  | **x** | ^2^ |
| **Cephalosporins and other beta-lactams** | | | | | | | | |
| Cefalexin | **x** | **x** |  |  |  |  |  | ^11,12^ |
| **Tetracyclines** | | | | | | | | |
| Doxycycline |  |  | **x** |  |  |  |  | ^2,11,12^ |
| **Aminoglycosides** | | | | | | | | |
| Gentamicin | **x** | **x** |  |  |  |  |  | ^11,12^ |
| **Macrolides** | | | | | | | | |
| Erythromycin |  |  | **x** |  |  |  |  | ^11,12^ |
| Clarithromycin |  |  | **x** | **x** |  |  | **x** | ^2,11,12^ |
| Azithromycin |  |  | **x** |  |  |  |  | ^11,12^ |
| **Fluoroquinolones** | | | | | | | | |
| Levofloxacin |  |  |  |  |  | **x** |  | ^15^ |
| Ciprofloxacin | **x** | **x** |  |  |  | **x** |  | ^11,12,15^ |
| **Sulfonamides and Trimethoprim** | | | | | | | | |
| Co-trimoxazole |  |  |  |  | **x** |  |  | ^13,16^ |

**Table S.2:** Mid-year estimates of the CPRD study population compared to Office of National Statistics (ONS) mid-year population estimates for England ^17^ for the study period from 2015-2018.

| **Year** | **2015** | **2016** | **2017** | **2018** |
| --- | --- | --- | --- | --- |
| **Mid-year population in CPRD study population** | | | | |
| 0-5m | 38,142 | 39,432 | 38,611 | 37,640 |
| 6-23 m | 238,809 | 240,573 | 242,893 | 239,744 |
| 2-4 y | 476,680 | 474,485 | 472,586 | 473,330 |
| 5-14 y | 1,399,998 | 1,453,645 | 1,503,173 | 1,537,273 |
| 15-44y | 5,019,852 | 5,133,276 | 5,274,704 | 5,410,860 |
| 45-64y | 3,161,115 | 3,239,387 | 3,313,310 | 3,368,794 |
| 65-74y | 1,129,375 | 1,160,644 | 1,183,455 | 1,199,898 |
| ≥75 y | 972,435 | 987,963 | 1,011,551 | 1,033,952 |
| Total | 12,436,406 | 12,729,405 | 13,040,283 | 13,301,311 |
| **ONS mid-year population England** | | | | |
| Total | 54,786,327 | 55,268,067 | 55,619,430 | 55,977, 178 |
| Proportion (%) | 22.7 | 23.03 | 23.45 | 23.76 |

Mid-year population estimates for the study population represent the number of patient IDs on 1^st^ July for each year of the study period. Proportion = proportion of the mid-year English population represented by the mid-year CPRD study population. Y= years, m = months.

**Equation S.1**: Initial model equation

$$E\left( y_{t}^{a} \right)= \beta_{0}^{a}P_{t}^{a}+\beta_{1}^{a}RSVP_{t}^{a}+\beta_{2}^{a}InfluenzaP_{t}^{a}+\beta_{3}^{a}AdenovirusP_{t}^{a}+ \beta_{4}^{a}hMPVP_{t}^{a}+ \beta_{5}^{a}ParainfluenzaP_{t}^{a}+\beta_{6}^{a}RhinovirusP_{t}^{a}+\beta_{7}^{a}S.pneumoniaeP_{t}^{a}+\beta_{8}^{a}M.pneumoniaeP_{t}^{a}+\beta_{9}^{a}temperatureP_{t}^{a}$$

The basic model is represented above where ($E\left( y_{t}^{a} \right)$) represents the count of expected outcomes (e.g., respiratory antibiotic prescriptions) for each age group (a) in the week (t). The right side of the equation contains the intercept ($\beta_{0}^{a})$, weekly counts of respiratory pathogens and average temperatures, each multiplied by the mid-year population for age group (a), in the week (t) ($P_{t}^{a}$).

**Table S.3:** Annual counts and rates of antibiotic and respiratory antibiotic prescriptions in CPRD and counts of laboratory-confirmed infections from SGSS over the study period from 2015-2018.

| **Year** | **2015** | **2016** | **2017** | **2018** | **Total** |
| --- | --- | --- | --- | --- | --- |
| Antibiotic prescriptions | 7,264,082 | 7,103,669 | 6,935,912 | 6,665,391 | 27,969,054 |
| Antibiotic prescriptions per 1000 | 584 | 558 | 532 | 501 | - |
| Respiratory antibiotic prescriptions | 3,566,383 | 3,488,283 | 3,292,600 | 3,083,950 | 13,431,216 |
| Respiratory antibiotic prescriptions per 1000 | 287 | 274 | 252 | 232 | - |
| Laboratory-confirmed respiratory infections | 33,602 | 42,784 | 44,755 | 71,797 | 192,938 |
| RSV | 8,123 | 8,200 | 8,449 | 11,408 | 36,180 |
| Influenza A | 3,464 | 8,538 | 7,844 | 16,347 | 36,193 |
| Influenza B | 1,410 | 3,120 | 2,362 | 14,194 | 21,086 |
| Adenovirus | 2,289 | 2,413 | 2,907 | 3,398 | 11,007 |
| Rhinovirus | 8,250 | 9,983 | 11,100 | 13,220 | 42,553 |
| hMPV | 1,244 | 1,453 | 1,933 | 2,696 | 7,326 |
| Parainfluenza | 3,212 | 2,989 | 4,202 | 4,366 | 14,769 |
| *S. pneumoniae* | 5,229 | 5,677 | 5,665 | 5,949 | 22,520 |
| *M. pneumoniae* | 381 | 411 | 293 | 219 | 1,304 |

**Table S.4:** Proportion of total prescriptions in CPRD for age groups by class and WHO AWaRe category^18^ for study period 2015-2018.

| Age | 0-5m N=46,850 | 6-23m N=760,701 | 2-4y N=1,135,558 | 5-14y N=1,851,882 | 15-44y N=7,761,032 | 45-64y N=6,844,164 | 65-74y N=4,088,064 | ≥75y N=5,480,803 | Total N=27,969,054 |
| --- | --- | --- | --- | --- | --- | --- | --- | --- | --- |
| **Class %** | | | | | | | | | |
| PEN | 77.6 | 81.5 | 75.9 | 67.0 | 46.2 | 43.0 | 40.1 | 38.7 | 46.7 |
| CEPH+ | 2.4 | 1.3 | 1.5 | 1.6 | 1.7 | 1.9 | 3.0 | 4.9 | 2.5 |
| TET | <0.1 | <0.1 | <0.1 | 3.8 | 15.6 | 15.6 | 15.0 | 11.0 | 12.8 |
| AMINO | 1.5 | 0.9 | 2.2 | 5.4 | 4.5 | 5.3 | 4.3 | 2.7 | 4.2 |
| MAC | 7.9 | 12.4 | 13.5 | 13.0 | 10.6 | 11.8 | 12.2 | 10.2 | 11.4 |
| CLI+ | <0.1 | <0.1 | <0.1 | <0.1 | 0.2 | 0.3 | 0.3 | 0.4 | 0.3 |
| OTHER | 0.1 | 0.1 | 0.1 | 0.2 | 0.2 | 0.4 | 0.5 | 0.3 | 0.3 |
| SULF+ | 9.8 | 3.3 | 6.0 | 7.1 | 7.0 | 7.7 | 9.3 | 14.0 | 8.7 |
| MTZ+ | 0.3 | 0.1 | 0.2 | 0.3 | 3.6 | 2.1 | 1.4 | 1.0 | 1.9 |
| QUIN | 0.1 | 0.2 | 0.2 | 0.4 | 1.7 | 2.4 | 2.6 | 2.5 | 2.0 |
| UTI | 0.2 | 0.2 | 0.4 | 1.1 | 8.7 | 9.5 | 11.2 | 14.3 | 9.3 |
| **AWaRe Category %** | | | | | | | | | |
| Access | 91.7 | 87.0 | 84.3 | 78.6 | 75.3 | 77.4 | 79.6 | 84.8 | 79.2 |
| Watch | 8.3 | 13.0 | 15.6 | 21.2 | 24.5 | 22.3 | 19.8 | 14.6 | 20.4 |
| Reserve | <0.1 | <0.1 | 0.1 | 0.2 | 0.1 | 0.1 | 0.2 | 0.1 | 0.1 |
| NA | <0.1 | <0.1 | <0.1 | <0.1 | 0.1 | 0.2 | 0.4 | 0.5 | 0.2 |

N = number of antibiotic prescriptions in CPRD over the study period. m = months, y = years, NA = no category, PEN = Penicillin's, CEPH+ = Cephalosporins & other beta lactams, TET = Tetracyclines, AMINO = Aminoglycosides, MAC = Macrolides, CLI+ = Clindamycin & Lincomycin, OTHER = Other antibacterials, SULF+ = Sulfonamides & Trimethoprim, MTZ+ = Metronidazole, Tinidazole & Ornidazole, QUIN = Quinolones, UTI = Urinary tract infection antibiotics.

**Figure S.1:** ACF plot comparing weekly laboratory-confirmed RSV infections in 5-14 years to 5-64 years. ACF plots measure the relationship between two time series, Xt and Yt.

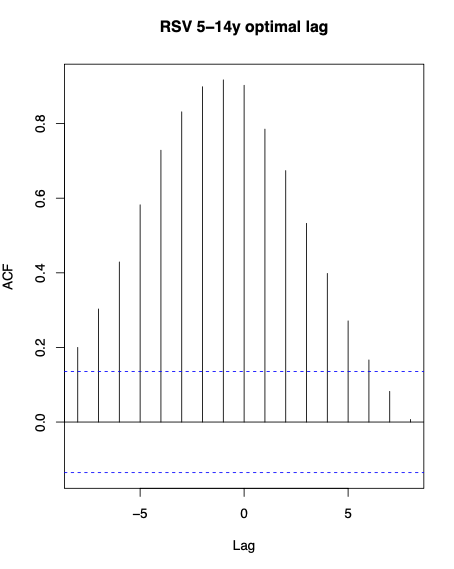

In this plot, Xt = time-series of weekly RSV infections for 5-14 years and Yt = time-series of weekly RSV infections for 5-64 years. The ACF plot demonstrates that RSV infections in 5-14 years peaks one week before RSV infections in 5-64 years (x is leading y).

**Figure S.2:** Correlation matrices of laboratory-confirmed respiratory infections for age groups 0-4 years (left), 5-64 years (middle), and ≥65 years (right), with average weekly temperatures over the study period.

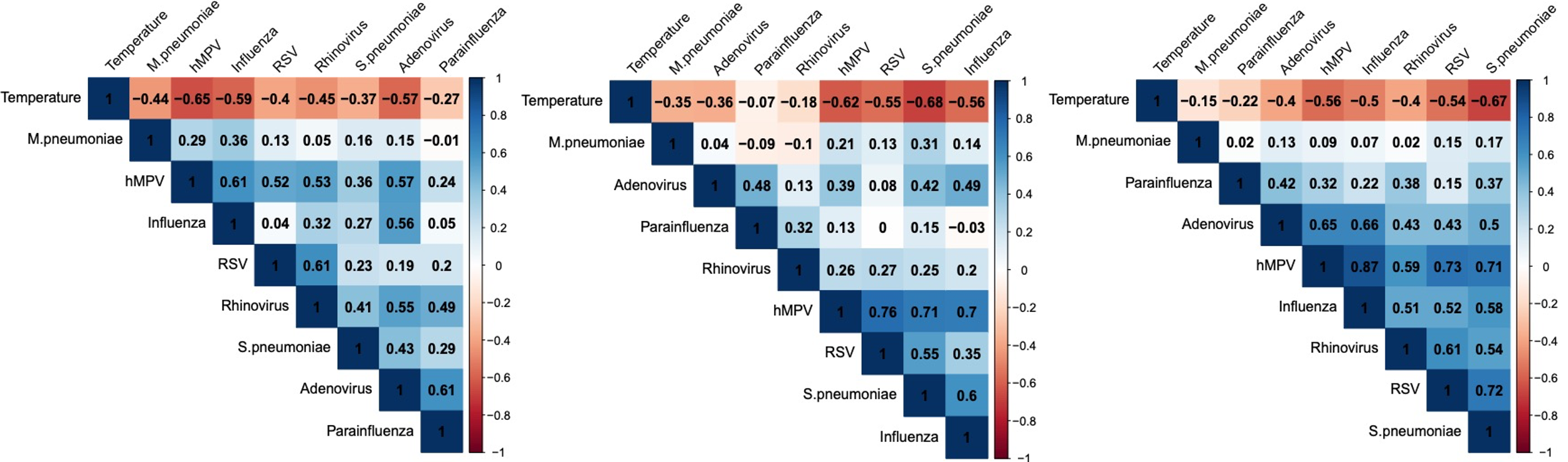

**Figure S.3: Development of age-specific models of respiratory antibiotic prescriptions according to Akaike Information Criterion (AIC) values.**

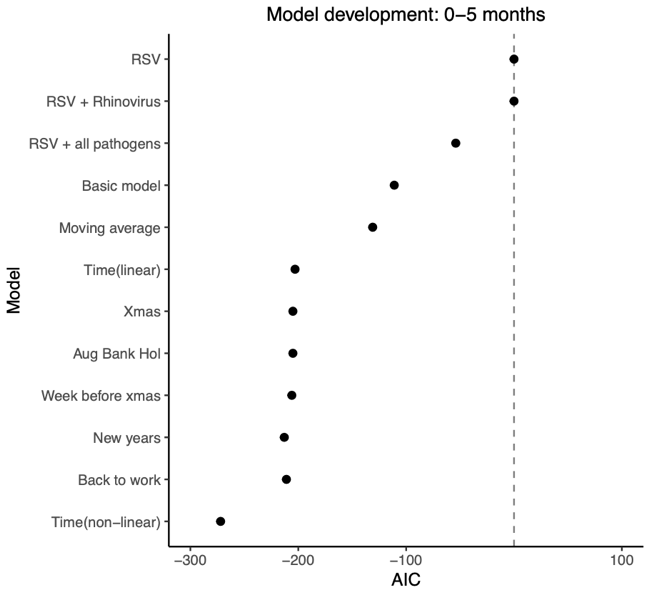

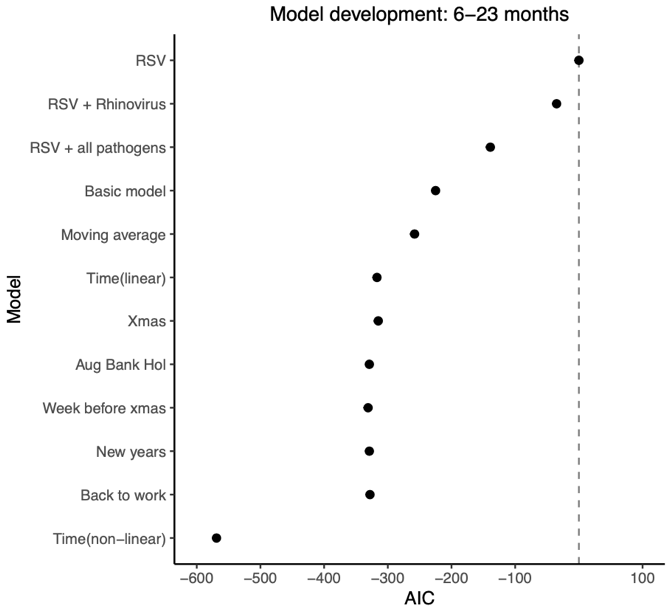

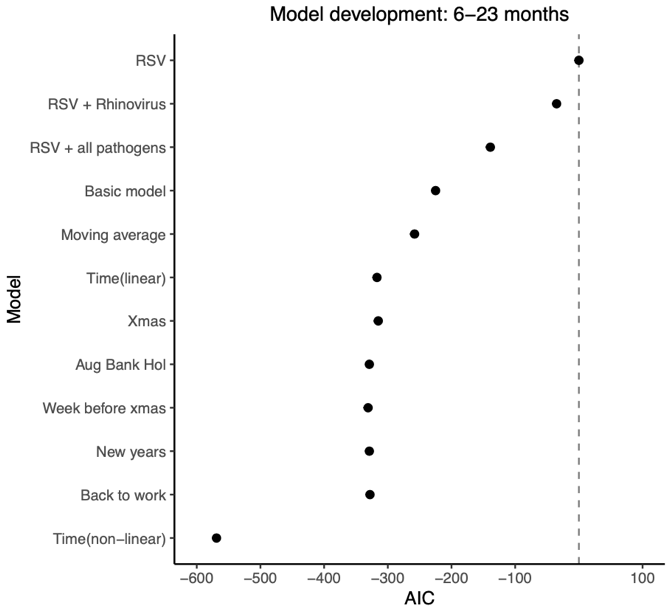

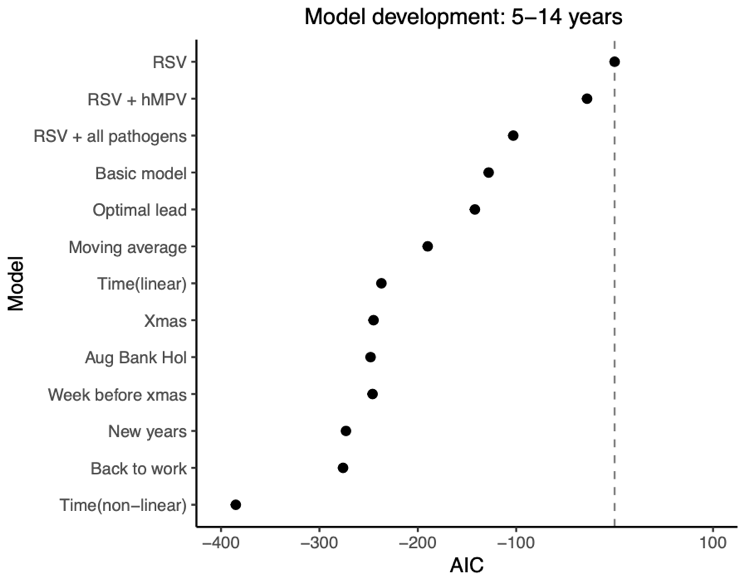

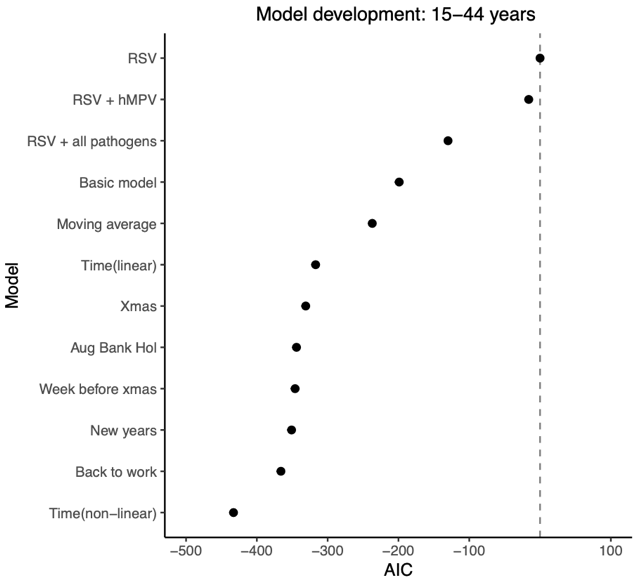

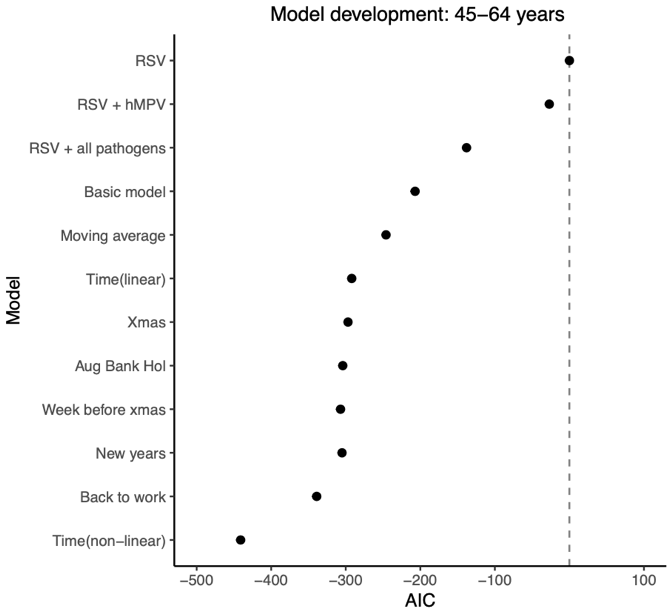

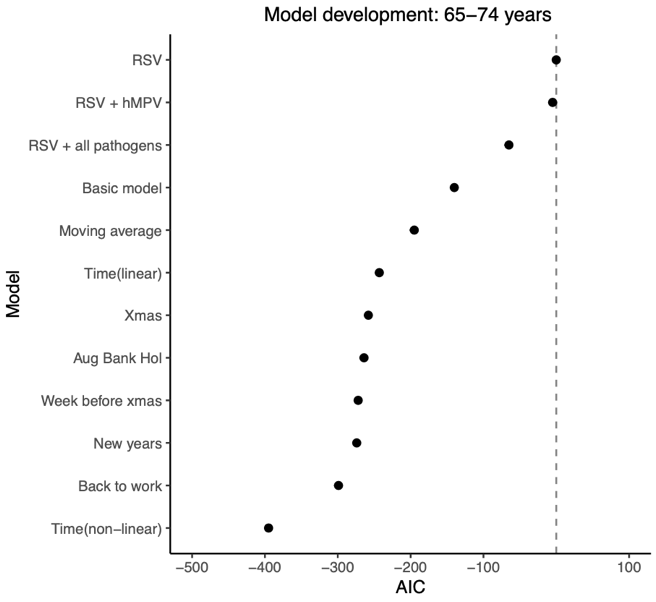

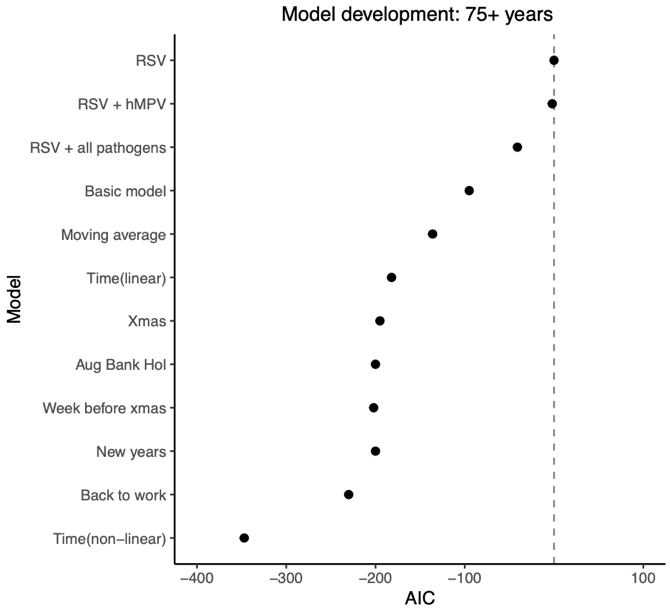

All AIC tests start with a model that includes only RSV and then evaluates the addition of the pathogen with the highest correlation with RSV in that age group (Figure S.2), as well as all pathogens. The basic model comprises of all exposure variables described in the initial model equation (Equation S.1). The moving average refers to the three-week moving average of laboratory-confirmed infections. Time (linear) represents a continuous variable for time (in weeks) to account for long-term linear trends in outcome counts not explained by exposures. Time (non-linear) adds a spline to control for long-term seasonal trends in outcome counts not explained by exposures. Indicator variables for practice holiday periods to adjust for outliers in antibiotic prescriptions include Xmas (Christmas holiday week), Aug Bank Hol (August Bank holiday week), New Years (week of New Year's), and back to work (week after New Year's).

Including a moving average for pathogen counts and outcome counts with a non-linear trend (spline) consistently provided the best or near-best fit across all age groups. Most indicators of practice closures had little impact on AIC for all age groups, except for the "back to work" week, which improved the model AIC for adults.

**Figure S.4:** An example of partial effect plots for the ≥75 years model of respiratory antibiotic prescriptions before truncating (Top) and after truncating (Below).

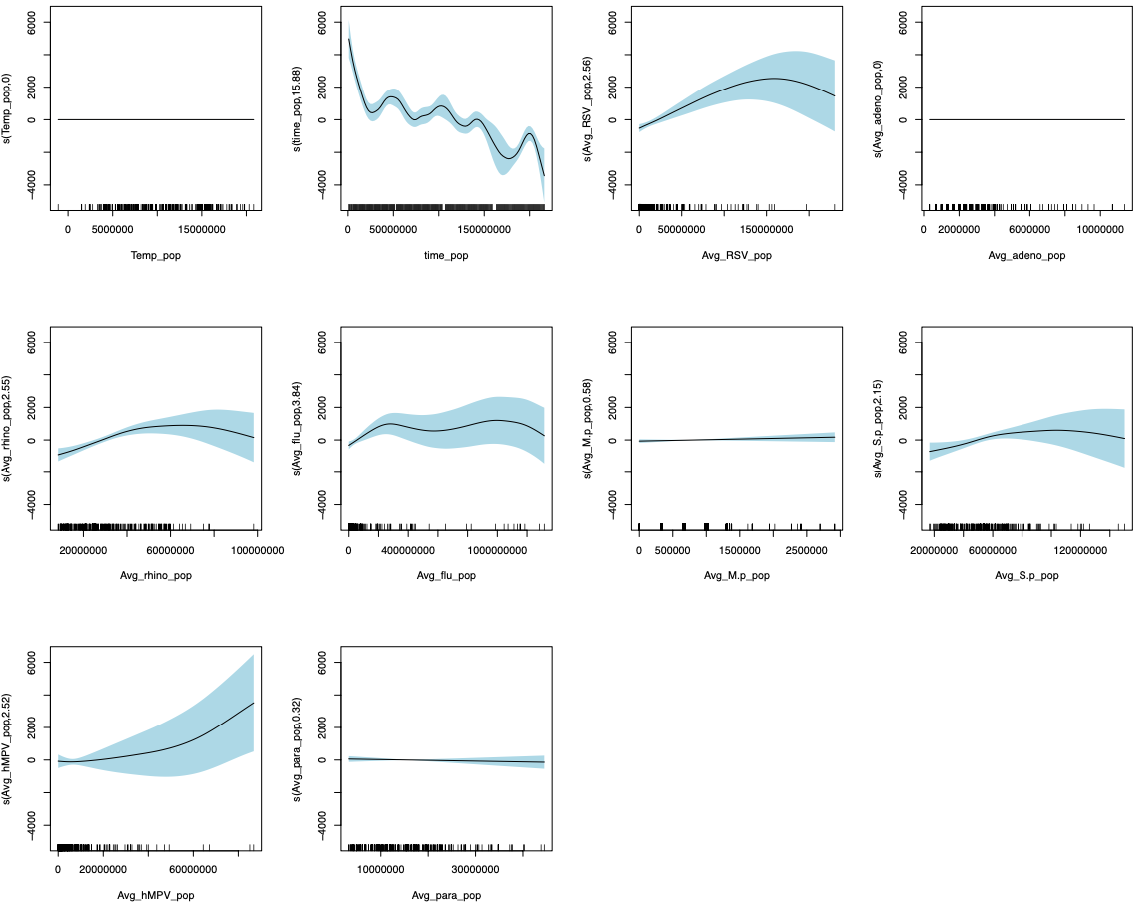

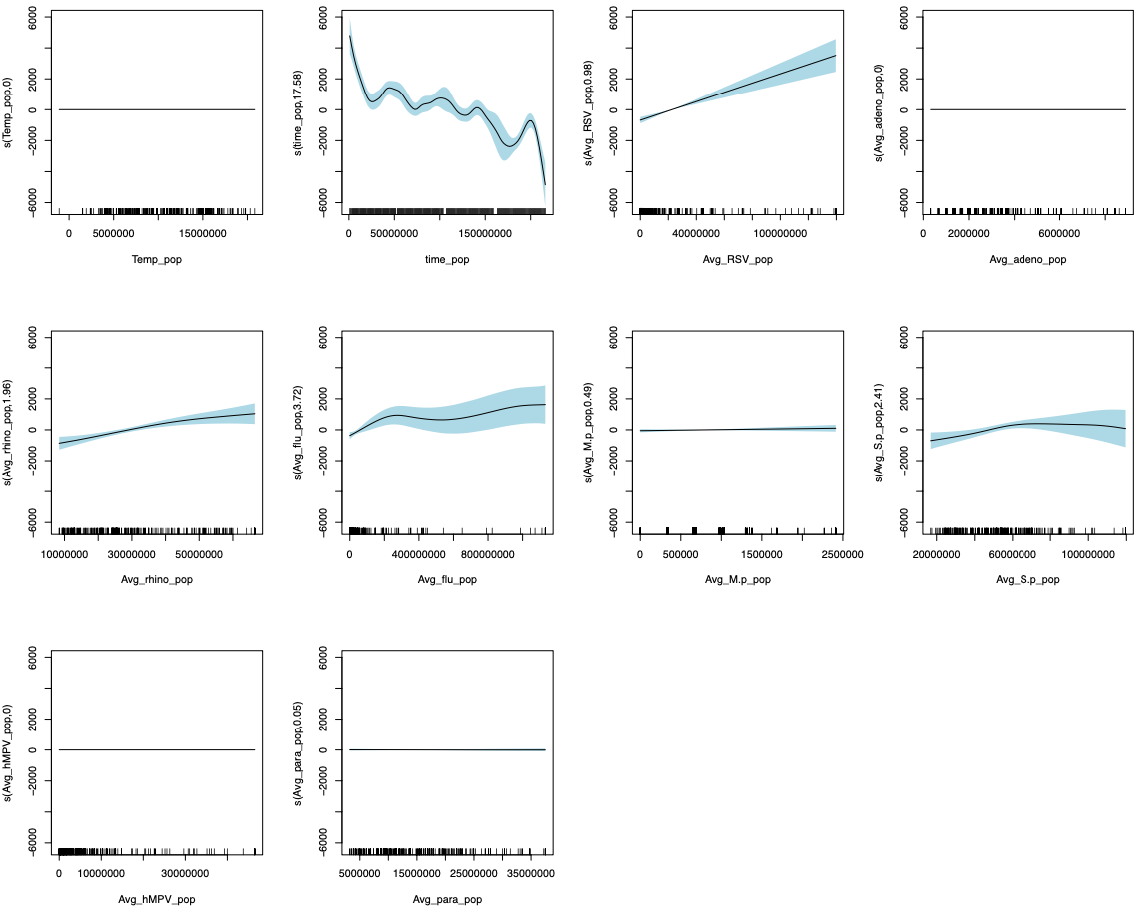

The X-axis is weekly counts of pathogens or average temperatures multiplied by the age-specific mid-year population (e.g. Avg_path_pop). The shaded blue area represents 95% confidence intervals of the predicted trend. A straight flat line (edf = 0) represents no relationship. Temp = temperature, adeno = adenovirus, rhino = rhinovirus, flu = influenza, M.p = *M. pneumoniae*, S.p = *S. pneumoniae*, para = parainfluenza.

**Figure S.5:** The final sets of time-varying explanatory covariates for all age-specific models demonstrated in the tables A-L below, inclusion indicated by an X.

| **A) Covariate** | **Respiratory antibiotics** | | | | | | | |
| --- | --- | --- | --- | --- | --- | --- | --- | --- |
|  | **0-5 m** | **6-23m** | **2-4 y** | **5-14 y** | **15-44y** | **45-64y** | **65-74y** | **≥75y** |
| **RSV** | x | x | x | x | x | x | x | x |
| **Influenza** | x | x | x | x | x | x | x | x |
| **Adenovirus** | x | x | x | x | x |  | x |  |
| **Rhinovirus** | x | x | x | x | x | x | x | x |
| **Parainfluenza** |  | x | x | x |  | x |  | x |
| **hMPV** | x |  | x | x |  | x |  |  |
| ***S. pneumoniae*** |  | x | x | x | x | x | x | x |
| ***M. pneumoniae*** |  | x |  | x | x | x |  | x |
| **Temperature** |  |  | x | x | x | x | x |  |

| **B) Covariate** | **Total antibiotics** | | | | | | | |
| --- | --- | --- | --- | --- | --- | --- | --- | --- |
|  | **0-5 m** | **6-23m** | **2-4 y** | **5-14 y** | **15-44y** | **45-64y** | **65-74y** | **≥75y** |
| **RSV** | x | x | x | x |  | x | x | x |
| **Influenza** |  | x | x | x |  | x |  |  |
| **Adenovirus** | x | x | x | x |  | x |  |  |
| **Rhinovirus** | x | x | x | x | x | x | x | x |
| **Parainfluenza** |  | x | x | x | x |  | x | x |
| **hMPV** |  |  | x | x |  |  |  |  |
| ***S. pneumoniae*** |  | x | x | x | x | x | x | x |
| ***M. pneumoniae*** |  | x |  | x | x | x |  | x |
| **Temperature** | x |  | x | x | x | x | x |  |

| **C) Covariate** | **Specific antibiotics potentially important for resistance** | | | | | | | |
| --- | --- | --- | --- | --- | --- | --- | --- | --- |
|  | **0-5 m** | **6-23m** | **2-4 y** | **5-14 y** | **15-44y** | **45-64y** | **65-74y** | **≥75y** |
| **RSV** | x | x | x | x |  | x | x | x |
| **Influenza** |  | x | x | x | x | x | x | x |
| **Adenovirus** | x | x | x | x | x | x | x |  |
| **Rhinovirus** | x | x | x | x | x | x | x | x |
| **Parainfluenza** |  | x | x | x | x |  | x | x |
| **hMPV** |  |  | x | x |  |  | x |  |
| ***S. pneumoniae*** |  | x | x | x | x | x | x | x |
| ***M. pneumoniae*** |  | x |  | x | x | x |  | x |
| **Temperature** | x |  | x | x | x |  |  |  |

| **D) Covariate** | **Nitrofurantoin prescriptions** | | | | | | | |
| --- | --- | --- | --- | --- | --- | --- | --- | --- |
|  | **0-5 m** | **6-23m** | **2-4 y** | **5-14 y** | **15-44y** | **45-64y** | **65-74y** | **≥75y** |
| **RSV** |  |  |  | x | x | x | x | x |
| **Influenza** |  |  |  | x | x | x | x | x |
| **Adenovirus** |  |  | x | x | x | x | x |  |
| **Rhinovirus** |  |  |  | x | x |  |  |  |
| **Parainfluenza** |  | x |  | x | x | x | x | x |
| **hMPV** |  |  |  | x | x | x | x | x |
| ***S. pneumoniae*** |  |  | x | x | x | x |  |  |
| ***M. pneumoniae*** |  |  | x | x |  |  |  | x |
| **Temperature** |  |  | x | x | x | x |  |  |

| **E) Covariate** | **Antibiotic classes 0-5m** | | | | | | | | | | |
| --- | --- | --- | --- | --- | --- | --- | --- | --- | --- | --- | --- |
|  | **Pen** | **Ceph** | **Tet** | **Amino** | **Mac** | **Clin** | **Other** | **Sulf** | **Met** | **Quin** | **UTI** |
| **RSV** | x |  |  |  | x |  |  | x |  |  |  |
| **Influenza** | x | x |  |  | x |  |  |  |  |  |  |
| **Adenovirus** | x | x |  |  | x |  |  | x |  |  |  |
| **Rhinovirus** | x |  |  |  | x |  |  |  |  |  |  |
| **Parainfluenza** |  |  |  |  |  |  |  | x |  |  |  |
| **hMPV** |  |  |  |  | x |  |  |  |  |  |  |
| ***S. pneumoniae*** |  |  |  |  |  |  |  | x |  |  |  |
| ***M. pneumoniae*** |  |  |  |  | x |  |  | x |  |  |  |
| **Temperature** |  |  |  |  | x |  |  |  |  |  |  |

| **F) Covariate** | **Antibiotic classes 6-23m** | | | | | | | | | | |
| --- | --- | --- | --- | --- | --- | --- | --- | --- | --- | --- | --- |
|  | **Pen** | **Ceph** | **Tet** | **Amino** | **Mac** | **Clin** | **Other** | **Sulf** | **Met** | **Quin** | **UTI** |
| **RSV** | x | x |  | x | x |  |  |  |  |  |  |
| **Influenza** | x | x |  | x | x |  |  |  |  | x |  |
| **Adenovirus** | x | x |  | x | x |  |  |  |  | x |  |
| **Rhinovirus** | x |  |  |  | x |  |  | x |  |  |  |
| **Parainfluenza** | x |  |  |  | x |  |  |  |  |  | x |
| **hMPV** |  |  |  |  | x |  |  |  |  |  |  |
| ***S. pneumoniae*** | x | x |  | x | x |  |  |  |  | x |  |
| ***M. pneumoniae*** | x | x |  |  |  |  |  |  |  |  |  |
| **Temperature** |  |  |  | x | x |  |  | x |  |  |  |
| **G) Covariate** | **Antibiotic classes 2-4y** | | | | | | | | | | |
|  | **Pen** | **Ceph** | **Tet** | **Amino** | **Mac** | **Clin** | **Other** | **Sulf** | **Met** | **Quin** | **UTI** |
| **RSV** | x | x |  |  | x |  | x | x |  | x |  |
| **Influenza** | x | x |  | x | x |  |  |  |  | x |  |
| **Adenovirus** | x |  |  | x | x |  |  |  |  | x | x |
| **Rhinovirus** | x | x |  | x | x |  | x | x |  |  |  |
| **Parainfluenza** | x |  |  |  | x |  |  | x |  |  |  |
| **hMPV** | x |  |  | x |  |  |  | x |  | x |  |
| ***S. pneumoniae*** | x |  |  | x | x |  |  | x | x |  | x |
| ***M. pneumoniae*** |  | x |  | x |  |  |  |  |  |  | x |
| **Temperature** | x | x |  | x |  |  | x | x |  | x | x |

| **H) Covariate** | **Antibiotic classes 5-14y** | | | | | | | | | | |
| --- | --- | --- | --- | --- | --- | --- | --- | --- | --- | --- | --- |
|  | **Pen** | **Ceph** | **Tet** | **Amino** | **Mac** | **Clin** | **Other** | **Sulf** | **Met** | **Quin** | **UTI** |
| **RSV** | x |  |  |  | x |  | x |  |  |  |  |
| **Influenza** | x | x | x | x | x |  |  | x |  |  | x |
| **Adenovirus** | x | x | x |  | x |  |  | x |  | x | x |
| **Rhinovirus** | x | x |  | x | x |  | x | x | x | x | x |
| **Parainfluenza** |  |  |  |  |  |  |  | x |  | x |  |
| **hMPV** | x |  | x |  | x |  |  | x |  |  |  |
| ***S. pneumoniae*** | x |  |  | x | x |  |  |  |  |  |  |
| ***M. pneumoniae*** | x |  | x |  |  |  |  |  |  |  | x |
| **Temperature** | x | x | x | x | x |  | x | x |  |  |  |

| **I) Covariate** | **Antibiotic classes 15-44y** | | | | | | | | | | |
| --- | --- | --- | --- | --- | --- | --- | --- | --- | --- | --- | --- |
|  | **Pen** | **Ceph** | **Tet** | **Amino** | **Mac** | **Clin** | **Other** | **Sulf** | **Met** | **Quin** | **UTI** |
| **RSV** |  |  |  | x |  |  |  |  | x |  | x |
| **Influenza** | x | x | x | x | x | x | x | x | x |  | x |
| **Adenovirus** | x | x |  | x |  |  | x |  | x | x | x |
| **Rhinovirus** | x | x | x |  | x |  |  | x | x |  | x |
| **Parainfluenza** | x | x | x |  | x | x |  | x | x | x | x |
| **hMPV** |  |  | x | x |  | x | x |  |  |  | x |
| ***S. pneumoniae*** | x | x |  | x | x | x |  | x | x | x | x |
| ***M. pneumoniae*** | x | x | x | x | x |  |  | x | x | x |  |
| **Temperature** | x | x | x |  | x |  |  | x | x | x | x |

| **J) Covariate** | **Antibiotic classes 45-64y** | | | | | | | | | | |
| --- | --- | --- | --- | --- | --- | --- | --- | --- | --- | --- | --- |
|  | **Pen** | **Ceph** | **Tet** | **Amino** | **Mac** | **Clin** | **Other** | **Sulf** | **Met** | **Quin** | **UTI** |
| **RSV** | x |  | x | x | x |  | x |  | x |  | x |
| **Influenza** | x | x | x | x | x | x | x |  | x |  | x |
| **Adenovirus** | x | x | x | x | x |  | x | x | x | x | x |
| **Rhinovirus** | x |  | x | x | x | x |  |  |  |  |  |
| **Parainfluenza** |  |  |  | x |  | x |  | x |  | x | x |
| **hMPV** |  |  | x | x |  | x |  |  |  | x | x |
| ***S. pneumoniae*** | x | x | x | x | x |  |  | x | x | x | x |
| ***M. pneumoniae*** | x | x | x | x | x |  | x | x |  | x |  |
| **Temperature** |  |  | x | x | x | x | x |  |  | x | x |

| **K) Covariate** | **Antibiotic classes 65-74y** | | | | | | | | | | |
| --- | --- | --- | --- | --- | --- | --- | --- | --- | --- | --- | --- |
|  | **Pen** | **Ceph** | **Tet** | **Amino** | **Mac** | **Clin** | **Other** | **Sulf** | **Met** | **Quin** | **UTI** |
| **RSV** | x | x | x | x | x |  | x | x | x |  | x |
| **Influenza** | x |  | x | x | x | x | x | x |  |  | x |
| **Adenovirus** | x |  |  |  | x |  | x |  | x |  |  |
| **Rhinovirus** | x | x | x |  | x | x |  |  |  |  |  |
| **Parainfluenza** | x |  | x | x |  |  |  | x | x | x | x |
| **hMPV** |  | x | x | x | x |  | x | x | x | x | x |
| ***S. pneumoniae*** | x |  | x | x | x |  |  |  | x |  |  |
| ***M. pneumoniae*** |  |  |  | x |  | x |  |  | x | x |  |
| **Temperature** | x | x | x | x |  | x |  | x |  | x |  |

| **L) Covariate** | **Antibiotic classes ≥75y** | | | | | | | | | | |
| --- | --- | --- | --- | --- | --- | --- | --- | --- | --- | --- | --- |
|  | **Pen** | **Ceph** | **Tet** | **Amino** | **Mac** | **Clin** | **Other** | **Sulf** | **Met** | **Quin** | **UTI** |
| **RSV** | x | x | x | x | x |  |  | x | x | x | x |
| **Influenza** |  |  | x | x | x |  | x |  |  |  | x |
| **Adenovirus** |  |  | x |  | x | x |  |  |  |  |  |
| **Rhinovirus** | x |  | x | x | x | x | x |  | x | x |  |
| **Parainfluenza** | x | x |  | x |  |  | x | x | x | x | x |
| **hMPV** |  | x |  | x | x |  |  | x |  | x | x |
| ***S. pneumoniae*** | x |  | x | x | x | x |  | x | x |  |  |
| ***M. pneumoniae*** | x |  | x | x |  | x |  | x | x |  | x |
| **Temperature** | x | x | x | x | x | x |  | x | x | x |  |

Tables E-L demonstrate explanatory time-changing covariates with evidence of a relationship with the outcome of interest for age-specific models of antibiotic classes. Class columns highlighted in red indicate that the model was not run because age-specific counts were <1,000 for the entire study period.

**Figure S.6:** Evidence of residual stationarity for age-specific models of respiratory antibiotic prescriptions, including ACF to test for autocorrelation and quantile-quantile plots (q-q) to test for normality of residuals.

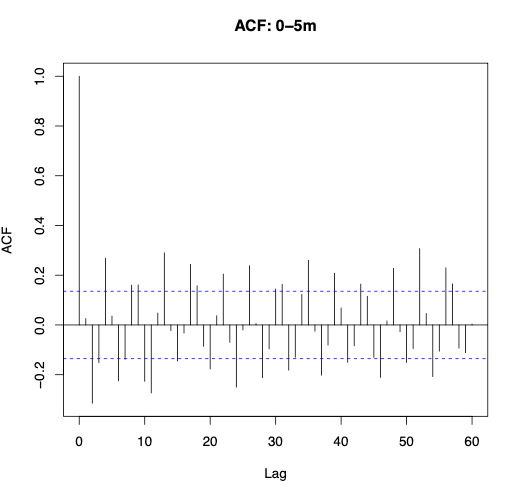

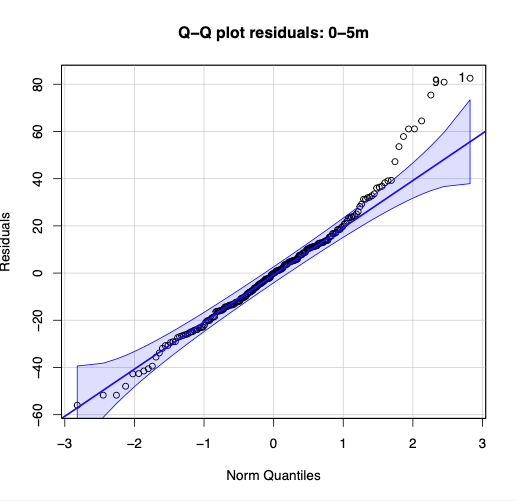

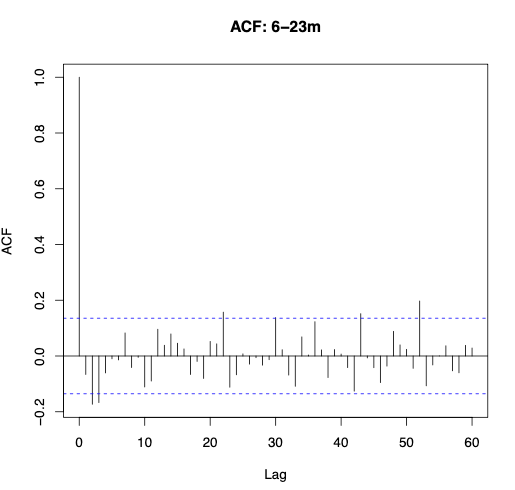

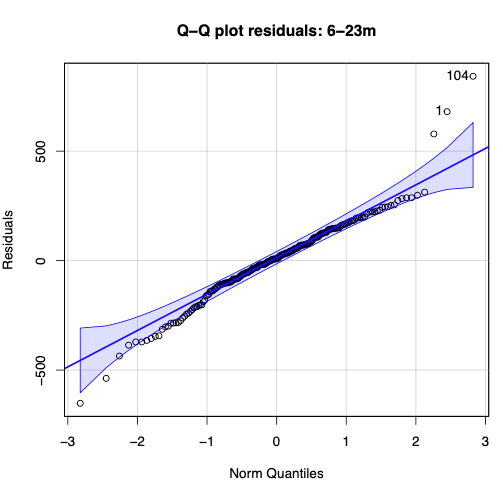

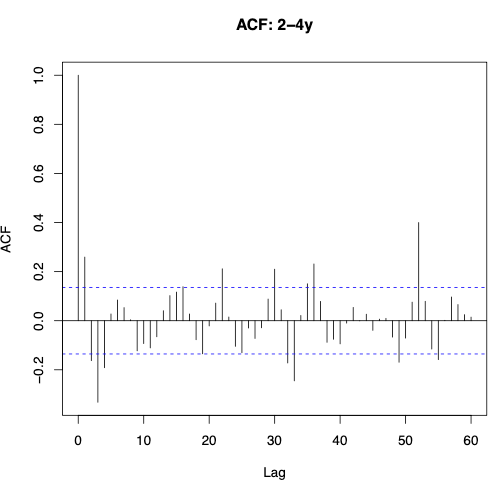

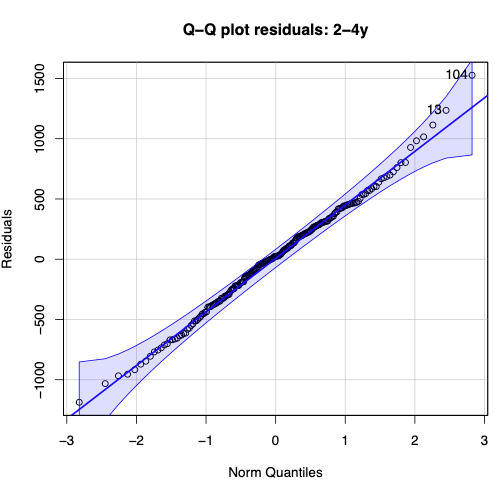

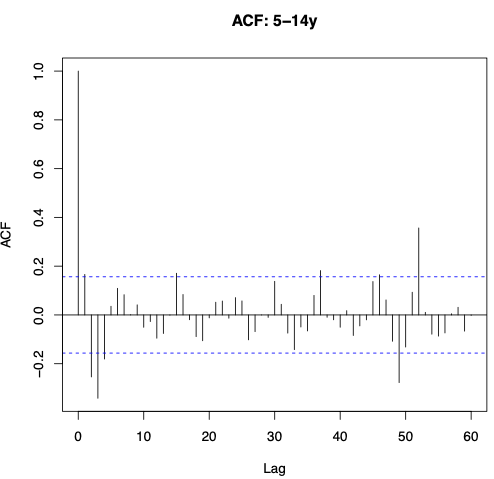

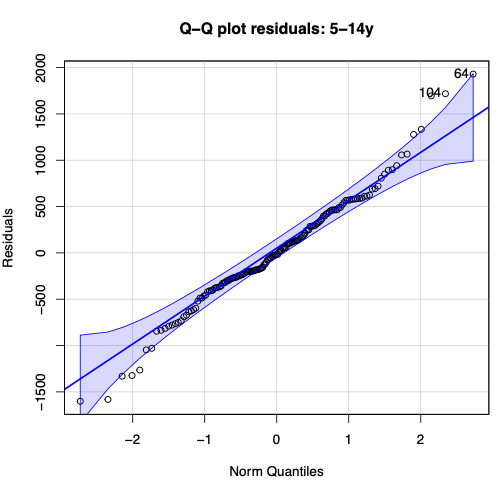

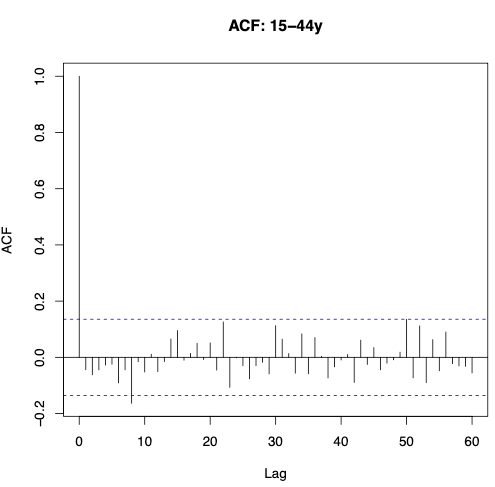

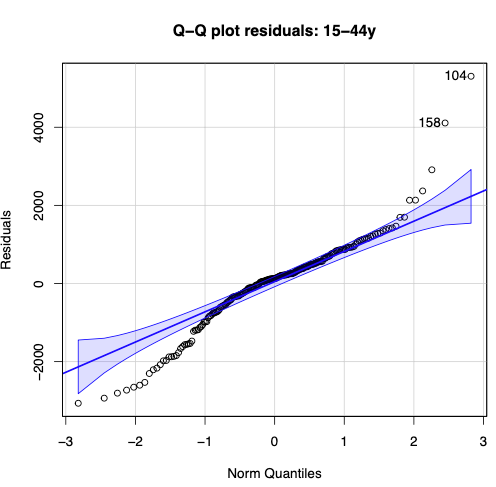

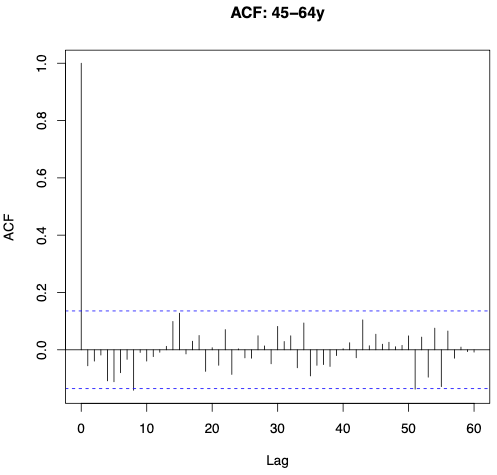

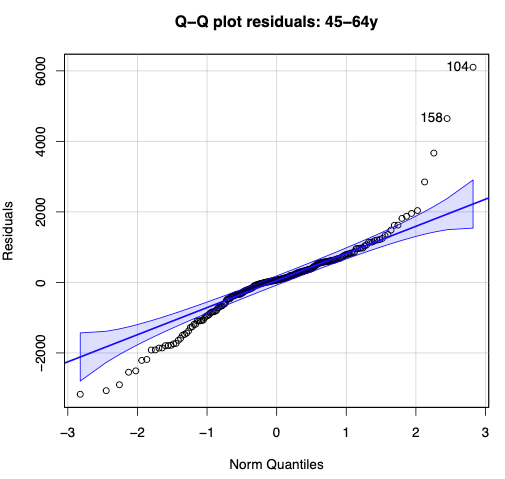

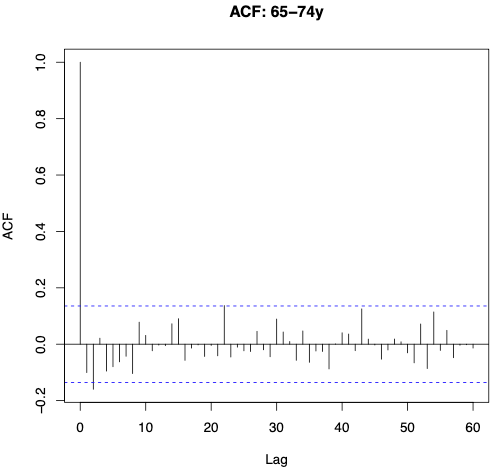

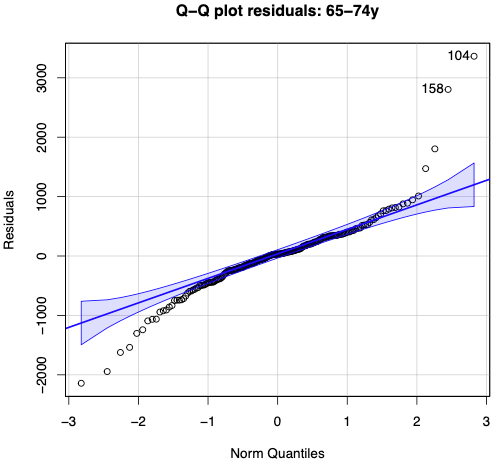

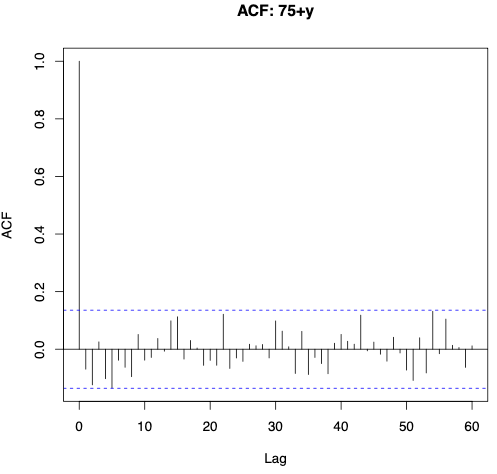

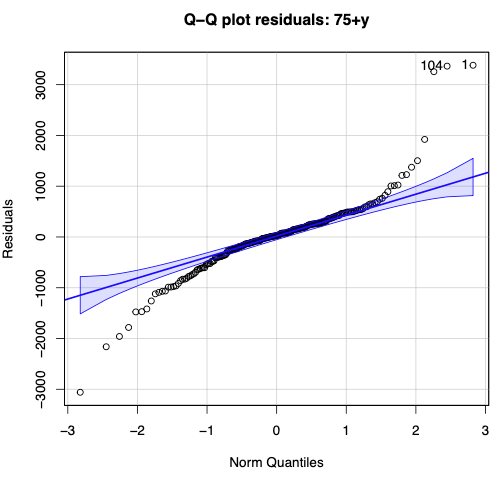

Overall, model residuals demonstrated reasonable normality with minimal heteroskedasticity or autocorrelation. ACF plots indicated potential autocorrelation for infants 0-5 months, children 2-4 years and 5-14 years. The potential autocorrelation for 0-5 months is likely due to unaccounted-for short-term prescription variations.^19^

**Figure S.7:** Attempts of autocorrelation adjustments for models of respiratory antibiotic prescriptions in 0-5 months, 2-4 years, and 5-14 years age groups

6-23 months

0-5 months

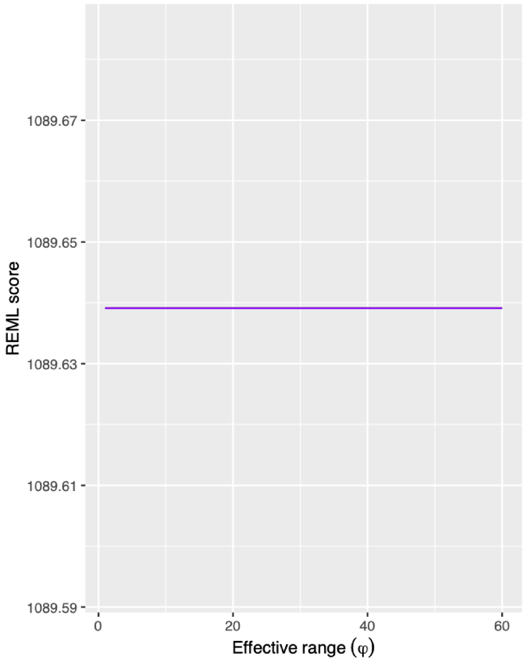

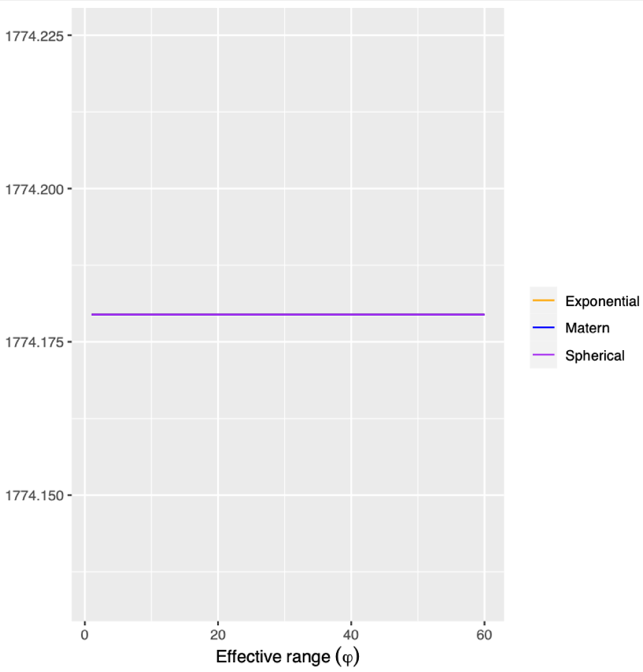

Mock data

5-14 years

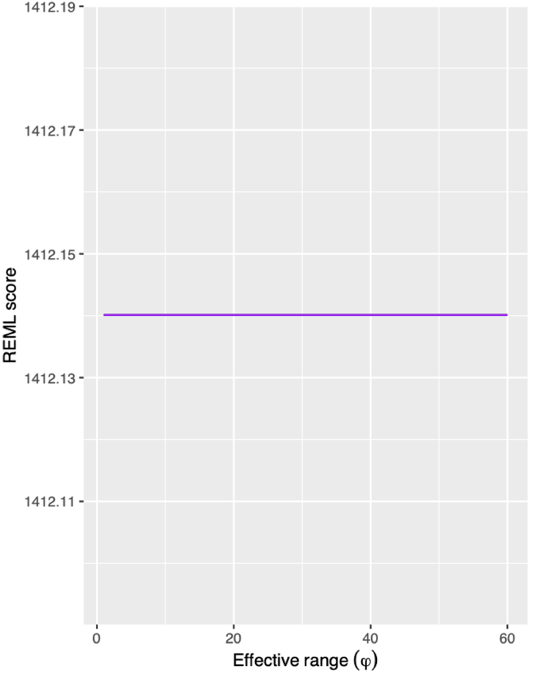

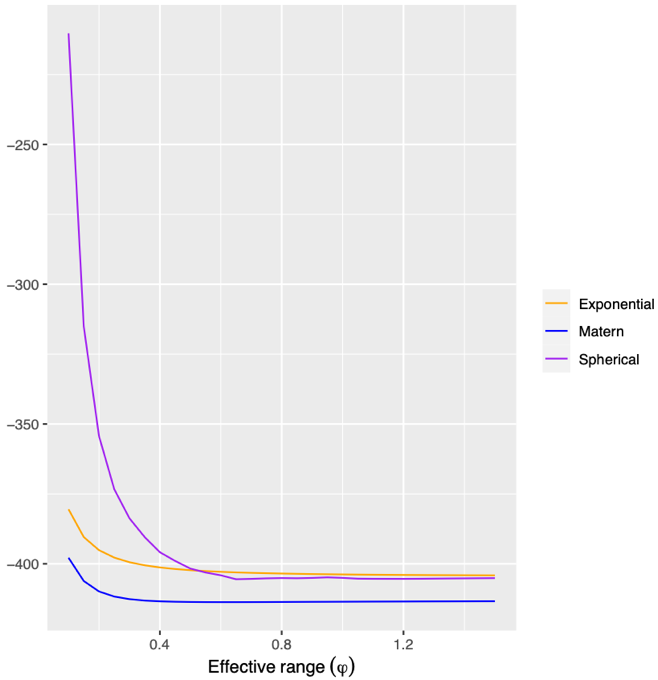

In GAMs, distinguishing between wiggly trends and autocorrelation is challenging.^20^ To adjust for autocorrelation, Gaussian process splines were applied following Simpson’s methodology.^20^ These splines use a penalised likelihood method similar to GAMS, but with an additional penalty from a correlation function. The first step involves identifying an effective range of Rho values (weekly lag) that provides the best REML score. GAMs were tested with weekly lags (1–60) using Matèrn, Squared Exponential, and Spherical correlation functions, and the REML score was extracted for each. The Rho value that minimised the REML score was identified through plotting. However, no change in REML was observed for any Rho value for all models. The Gaussian process spline method was validated with a mock dataset from stack exchange^21^ producing similar Rho values that minimised the spherical correlation function's REML score (around 0.65).

**Figure S.8:** Average model posterior simulations of respiratory antibiotic prescriptions compared to observations for scenarios where RSV is present and absent for age groups between 0-4 years over the study period.

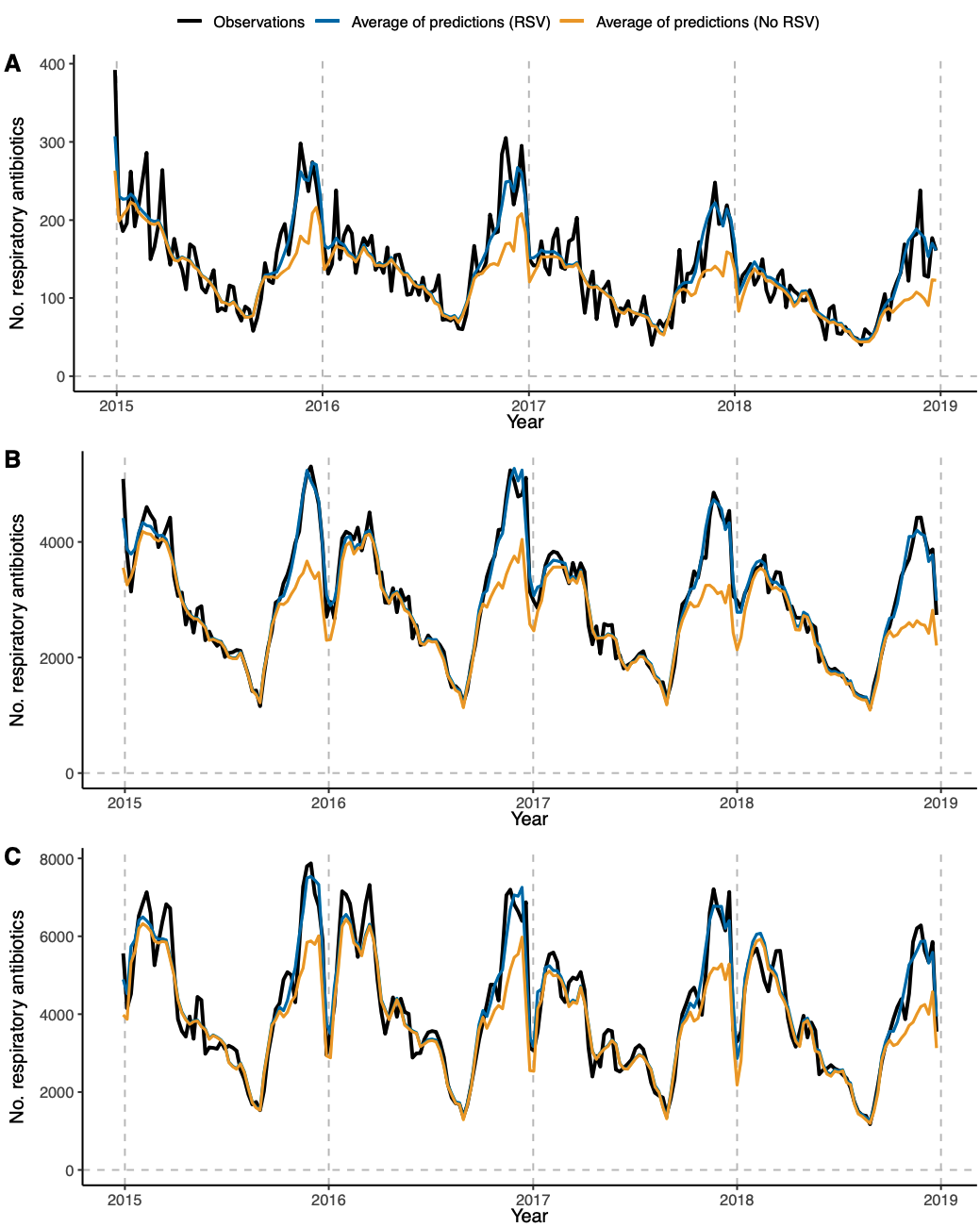

Black line = observational counts of weekly respiratory antibiotic prescriptions, blue line = average posterior simulations of weekly respiratory antibiotic prescriptions for a scenario where RSV is present, orange line = average posterior simulations of weekly respiratory antibiotic prescriptions for a scenario where RSV is absent. A = 0-5 months, B = 6-23 months, C = 2-4 years.

**Figure S.9:** Average model posterior simulations of respiratory antibiotic prescriptions compared to observations for scenarios where RSV is present and absent for age groups between 5-64 years over the study period.

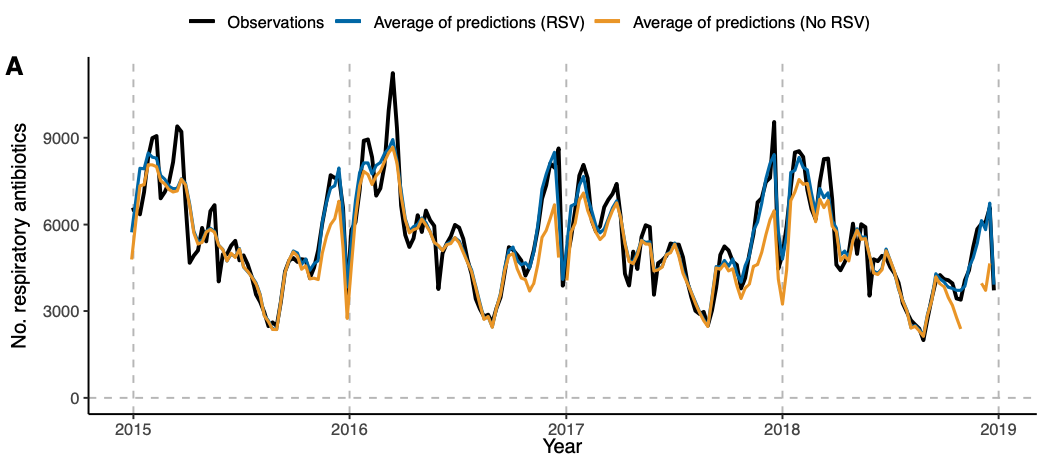

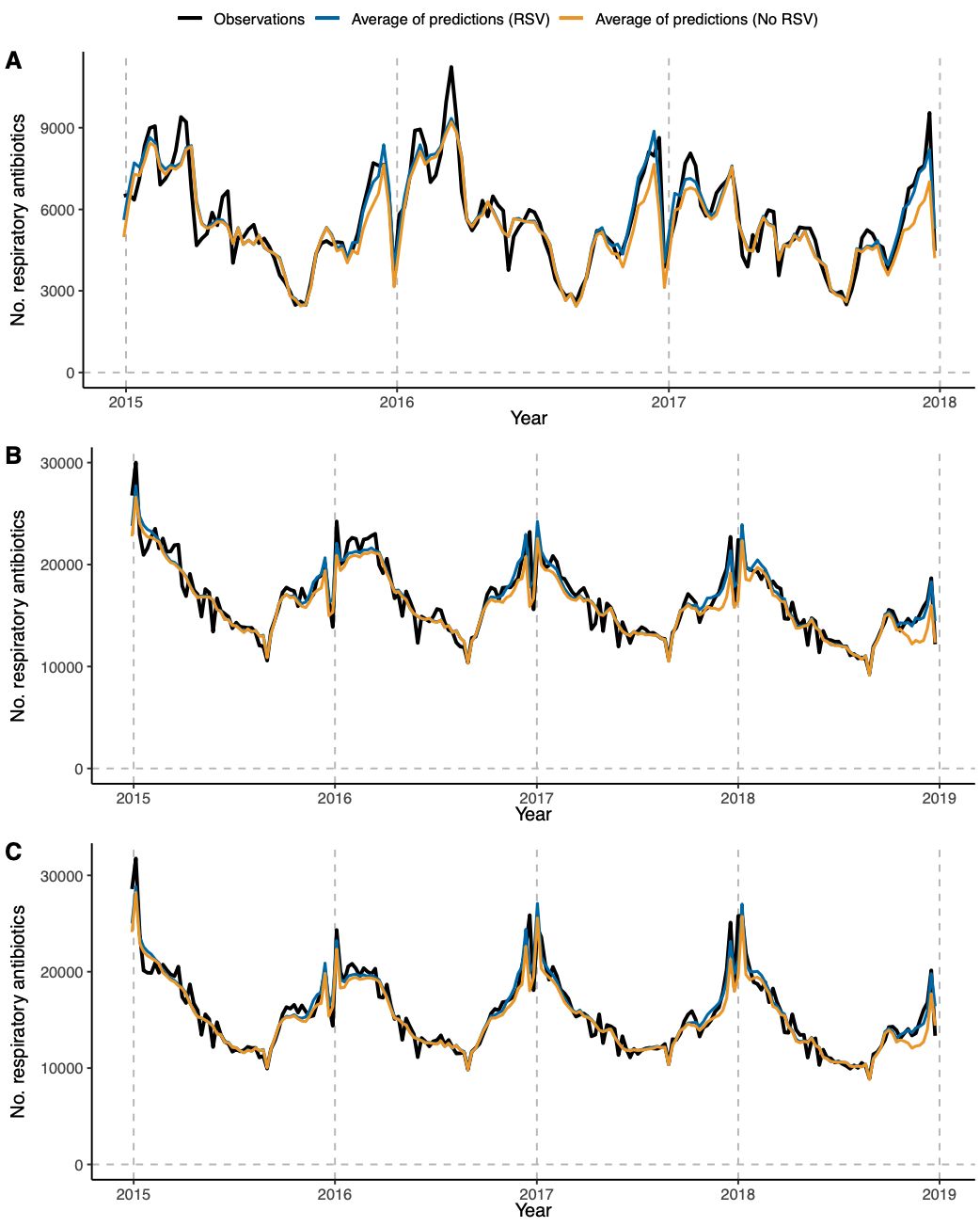

Black line = observational counts of weekly respiratory antibiotic prescriptions, blue line = average posterior simulations of weekly respiratory antibiotic prescriptions for a scenario where RSV is present, orange line = average posterior simulations of weekly respiratory antibiotic prescriptions for a scenario where RSV is absent. A = 5-14 years, B = 15-44 years, C = 45-64 years.

**Figure S.10:** Average model posterior simulations of respiratory antibiotic prescriptions compared to observations for scenarios where RSV is present and absent for age groups ≥65 years over the study period.

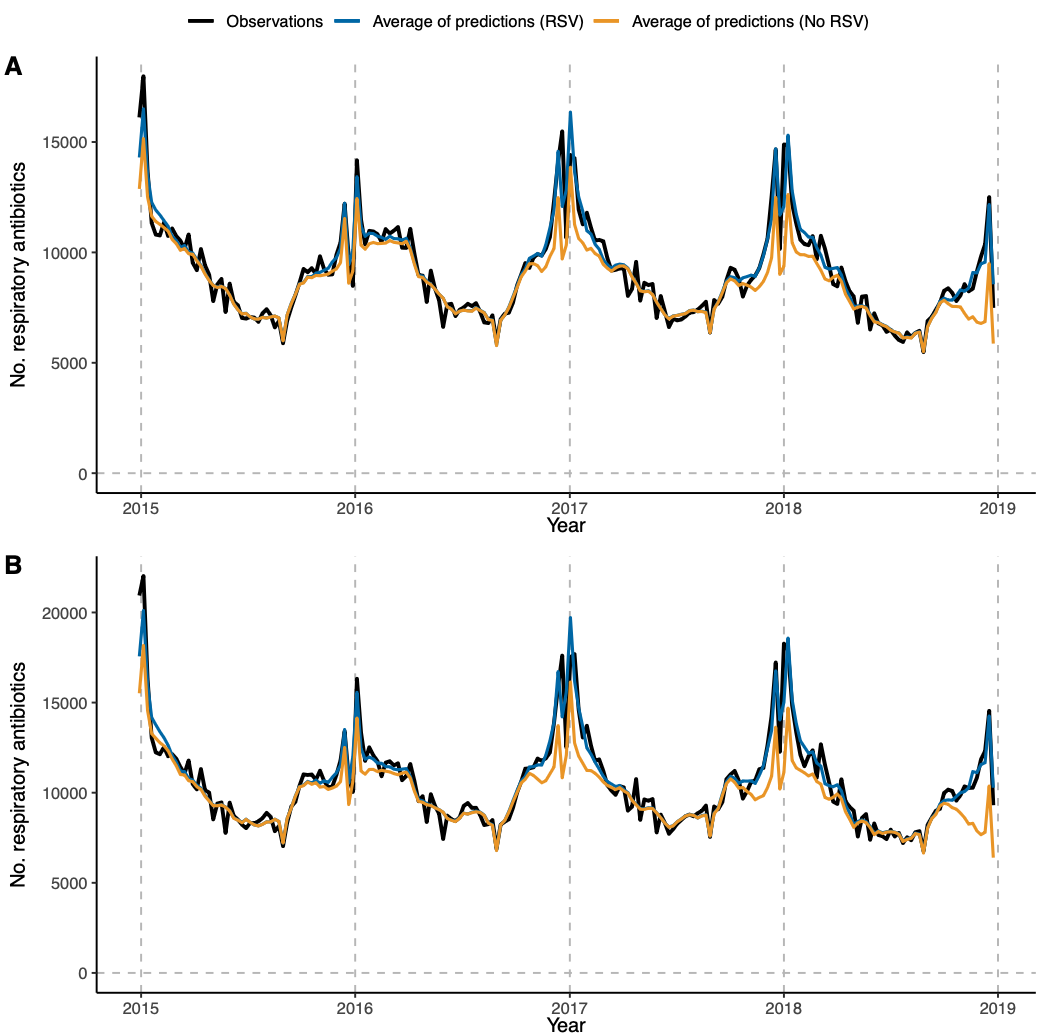

Black line = observational counts of weekly respiratory antibiotic prescriptions, blue line = average posterior simulations of weekly respiratory antibiotic prescriptions for a scenario where RSV is present, orange line = average posterior simulations of weekly respiratory antibiotic prescriptions for a scenario where RSV is absent. A = 65-74 years, B = ≥75 years.

**Figure S.11:** Average model posterior simulations of respiratory antibiotic prescriptions compared to observations for scenarios where RSV is present and absent for alternative 5–14 years models.

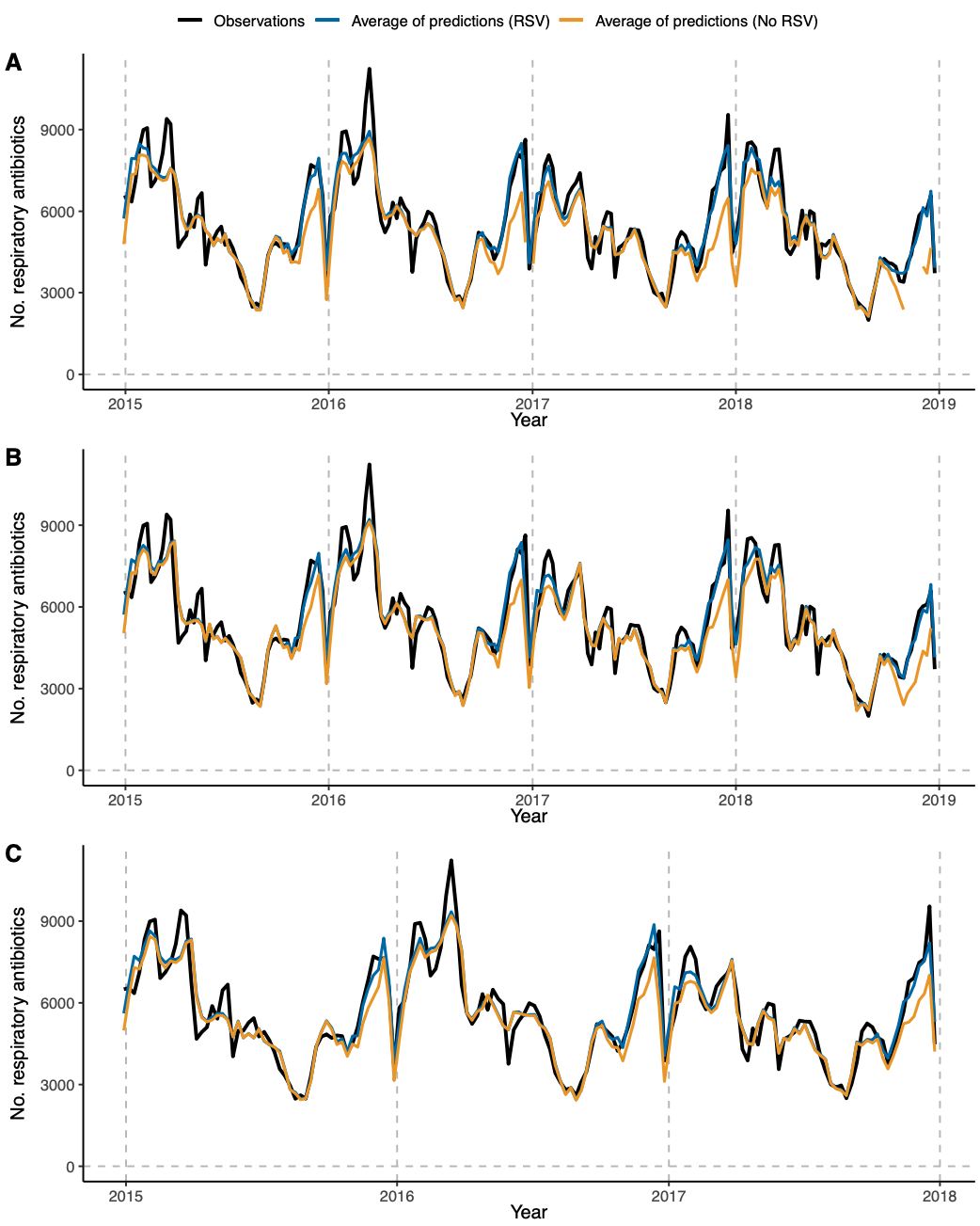

Black line = observational counts of weekly respiratory antibiotic prescriptions, blue line = average posterior simulations of weekly respiratory antibiotic prescriptions for a scenario where RSV is present, orange line = average posterior simulations of weekly respiratory antibiotic prescriptions for a scenario where RSV is absent. A = original 5–14 year model, B = 5–14 years model with seasonality trends decomposed, C = 5–14 years model with seasonality trends decomposed and counts of the final 2018 year removed.

**Table S.5:** Average annual RSV-attributable GP antibiotic prescriptions and respiratory antibiotic prescriptions potentially important for resistance compared to RSV-attributable GP respiratory antibiotic prescriptions from 29 December 2014 to 30 December 2018, stratified by age.

|  | **Respiratory antibiotic prescriptions (primary outcome)** | | | | **Total antibiotic prescriptions** | | | | **Respiratory antibiotic prescriptions potentially important for resistance** | | | |
| --- | --- | --- | --- | --- | --- | --- | --- | --- | --- | --- | --- | --- |
| **Age** | **Prescriptions**  **(95% CrI)** | **%** | **AP % (95% CrI)** | **Rate per 100,000**  **(95% CrI)** | **Prescriptions**  **(95% CrI)** | **%** | **AP % (95% CrI)** | **Rate per 100,000**  **(95% CrI)** | **Prescriptions**  **(95% CrI)** | **%** | **AP % (95% CrI)** | **Rate per 100,000**  **(95% CrI)** |
| **0-5 m** | 6,880*  (3,211-10,167) | 1.1 | 11  (5-17) | 2,100  (984-3,100) | 7,395*  (3,554-10,944) | 1.1 | 7  (4-11) | 2,258  (1,084-3,333) | 6,997*  (3,432-10,408) | 1.1 | 8  (4-12) | 2,136  (1,050-3,180) |
| **6-23 m** | 65,535*  (45,035-86,144) | 10.2 | 10  (7-13) | 6,580  (4,522-8,651) | 66,844*  (44,408-88,939) | 10.1 | 8  (6-11) | 6,711  (4,462-8,933) | 66,524*  (43,206-89,542) | 10.1 | 9  (6-12) | 6,679  (4,346-8,994) |
| **2-4 y** | 72,500  (29,222-119,821) | 11.3 | 8  (3-13) | 3,497  (1,405-5,782) | 75,227  (25,594-126,977) | 11.3 | 6  (2-10) | 3,628  (1,250-6,132) | 74,221  (24,484-125,501) | 11.3 | 7  (2-11) | 3,580  (1,179-6,054) |
| **5-14 y #** | 55,860  (-41,759-156,160) | 8.7 | 4  (-3-12) | 857  (-637-2,397) | 62,369  (-74,815-193,426) | 9.4 | 3  (-4-9) | 956  (-1,149-2,966) | 61,682  (-59,192-172,309) | 9.4 | 4  (-3-10) | 946  (-902-2,649) |
| **15-44 y** | 95,554  (6,792-185,445) | 14.9 | 3  (0-5) | 447  (31-868) | - | - | - | - | - | - | - | - |
| **45-64 y** | 86,608  (-16,608-191,877) | 13.5 | 2  (0-6) | 612  (-117-1,357) | 136,809  (3,015-273,393) | 20.6 | 2  (0-4) | 966  (18-1,934) | 148,091  (45,216-253,682) | 22.5 | 3  (1-5) | 1,046  (319-1,790) |
| **65-74 y** | 107,893  (60,407-158,507) | 16.9 | 5  (3-7) | 1,972  (1,104-2,901) | 123,365  (34,494-212,128) | 18.6 | 3  (1-4) | 2,255  (608-3,877) | 138,206  (65,507-207,870) | 21.0 | 4  (2-6) | 2,526  (1,185-3,804) |
| **≥75 y** | 149,078  (93,733-206,045) | 23.3 | 6  (4-8) | 3,279  (2,050-4,532) | 192,246  (89,342-300,243) | 28.9 | 3  (1-5) | 4,229  (1,962-6,617) | 163,106  (85,334-246,094) | 24.8 | 4  (2-6) | 3,588  (1,872-5,415) |

AP = Attributable Proportion, % = age-specific proportion, CrI = credible interval, m = months, y = years. *= Prescriptions are estimated assuming English mid-year populations are equally distributed by month of age as ONS population estimates are only provided by year of age, - = The model was not run as age-specific counts of antibiotics demonstrated no relationship with confirmed RSV infections, # = Average annual RSV-attributable prescriptions for 5-14 years were estimated from 2015 to 2017 and assumed to apply to 2018, as 2018 data was excluded for this group (see model fitting).

**Figure S.12:** Age-specific class proportion % of RSV-attributable GP antibiotic prescriptions (Table A) and respiratory antibiotic prescriptions (Table B).

| **A** | **Class proportion of age-specific RSV-attributable GP antibiotic prescriptions %** | | | | | | | | | |
| --- | --- | --- | --- | --- | --- | --- | --- | --- | --- | --- |
| **Age** | **PEN** | **CEPH+** | **TET** | **AMINO** | **MAC** | **CLI+** | **OTHER** | **SULF+** | **MTZ+** | **QUIN** |
| **0-5 m** | 100 |  |  |  |  |  |  |  |  |  |
| **6-23 m** | 89 |  |  |  | 11 |  |  |  |  |  |
| **2-4 y** | 84 | 1 |  |  | 15 |  |  |  |  |  |
| **5-14 y** |  |  |  |  |  |  |  |  |  |  |
| **15-44 y** |  |  |  |  |  |  |  |  |  |  |
| **45-64 y** | 65 |  | 21 |  | 13 |  |  |  |  |  |
| **65-74 y** | 45 | 3 | 25 | 5 | 19 |  | 1 | 6 |  |  |
| **≥75 y** | 49 | 3 | 19 | 3 | 15 |  |  | 9 |  | 2 |

| **B** | **Class proportion of age-specific RSV-attributable GP respiratory antibiotic prescriptions %** | | |
| --- | --- | --- | --- |
| **Age** | **Penicillins** | **Tetracyclines** | **Macrolides** |
| **0-5 m** | 100 |  |  |
| **6-23 m** | 89.3 |  | 10.7 |
| **2-4 y** | 84.9 |  | 15.1 |
| **5-14 y** |  |  |  |
| **15-44 y** |  |  |  |
| **45-64 y** | 65.4 | 21.3 | 13.3 |
| **65-74 y** | 52.4 | 27.2 | 20.4 |
| **≥75y** | 59.0 | 22.9 | 18.1 |

m = months, y = years, PEN = Penicillin's, CEPH+ = Cephalosporins & other beta lactams, TET = Tetracyclines, AMINO = Aminoglycosides, MAC = Macrolides, CLI+ = Clindamycin & Lincomycin, OTHER = Other antibacterials, SULF+ = Sulfonamides & Trimethoprim, MTZ+ = Metronidazole, Tinidazole & Ornidazole, QUIN = Quinolones. UTI antibiotics were excluded from class proportion estimates of RSV attributable antibiotic prescriptions as these are assumed to not be prescribed for RSV infections.

**Table S.6:** Average annual RSV-attributable GP nitrofurantoin prescriptions from 29 December 2014 to 30 December 2018, stratified by age

| **Nitrofurantoin prescriptions (negative control)** | | | | |
| --- | --- | --- | --- | --- |
| **Age** | **Prescriptions**  **(95% CrI)** | **%** | **Attributable proportion % (95% CrI)** | **Rate per 100,000**  **(95% CrI)** |
| **0-5 m** | - | - | - | - |
| **6-23 m** | - | - | - | - |
| **2-4 y** | -33 (-324-266) | NA | -1 (-7-6) | -2 (-16-13) |
| **5-14 y** | - | - | - | - |
| **15-44 y** | 24,344 (6,588-45,611) | 32.0 | 4 (1-7) | 144 (31-214) |
| **45-64 y** | 18,232 (1,296-35,474) | 24.0 | 3 (0-5) | 129 (9-251) |
| **65-74 y** | 12,069 (1,055-22,151) | 15.9 | 2 (0-4) | 221 (18-404) |
| **≥75 y** | 21,380 (5,083-37,282) | 28.1 | 3 (1-4) | 470 (110-817) |

% = age-specific proportion, CrI = credible interval, m = months, y = years, - = The model was not run because age-specific counts of nitrofurantoin prescriptions were <1,000 during the study period or demonstrated no relationship with confirmed RSV infections. N/A = negative result not included in proportion estimate.

**Figure S.13**: Posterior simulations of weekly nitrofurantoin prescriptions for 15-44 years (top) and ≥75 years (bottom) for scenarios where RSV is present and absent from 2015 to 2018.

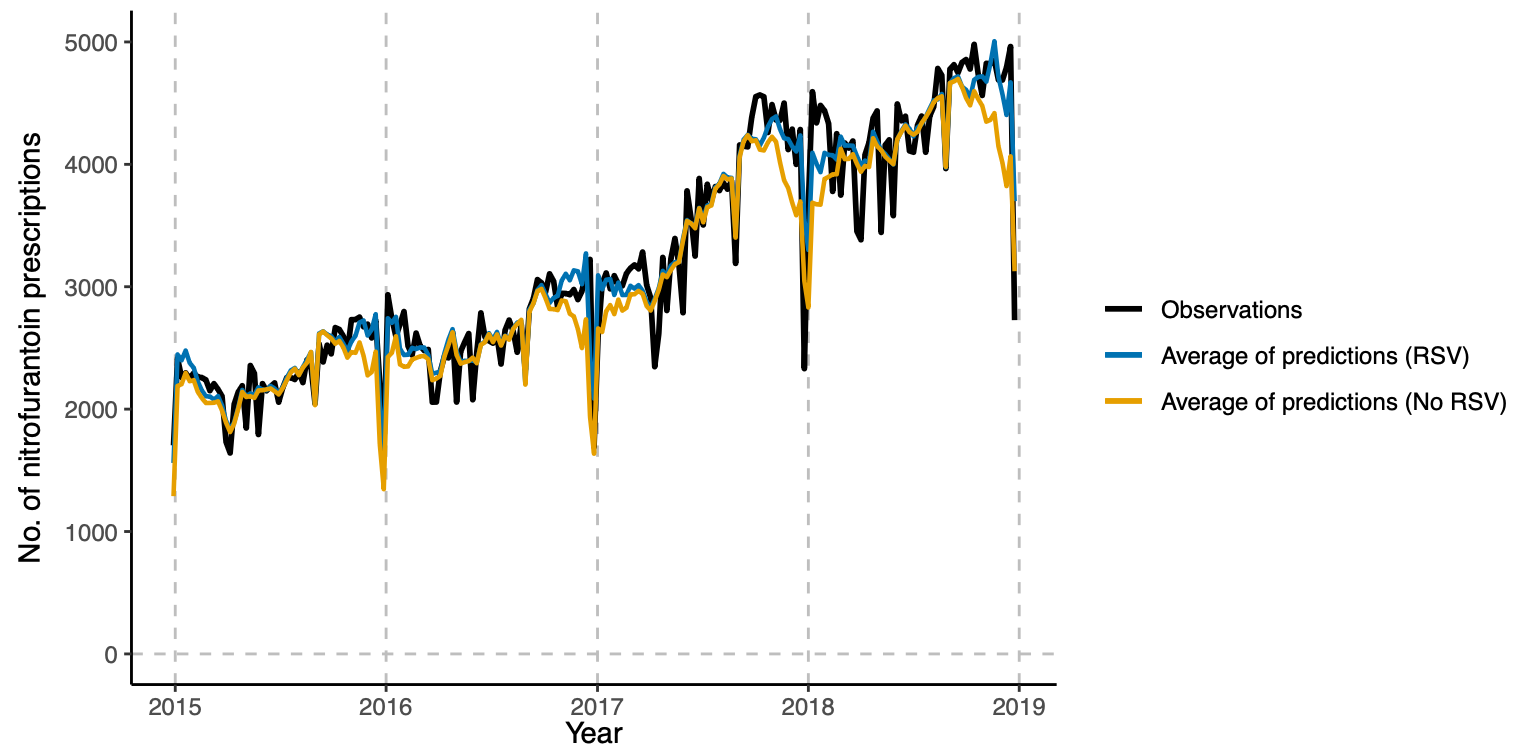

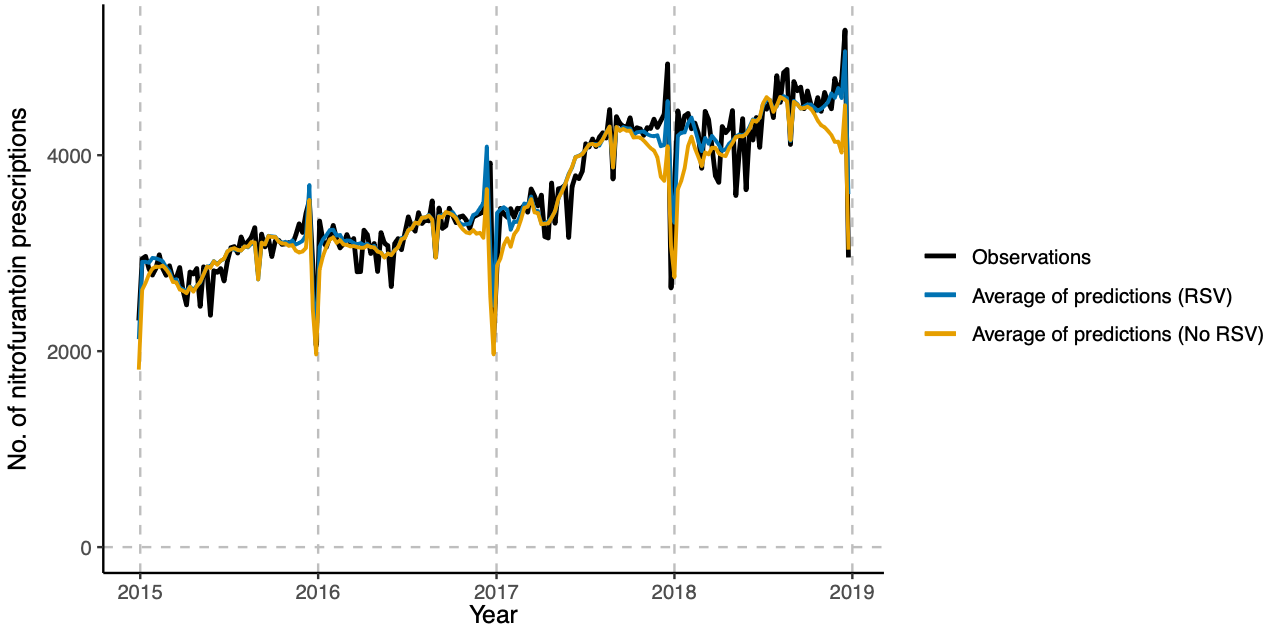

**Table S.7:** Average annual RSV-attributable GP respiratory antibiotic prescriptions with removal of highly correlated pathogens (middle) and removal of all pathogens except for RSV and Influenza (right) compared to primary outcome from 29 December 2014 to 30 December 2018, stratified by age.

|  | **Primary outcome** | | | **Removal of pathogens highly correlated with RSV** | | | **Only controlling for RSV and Influenza** | | |
| --- | --- | --- | --- | --- | --- | --- | --- | --- | --- |
| **Age** | **Prescriptions**  **(95% CrI)** | **AP %**  **(95% CrI)** | **Rate per 100,000**  **(95% CrI)** | **Prescriptions**  **(95% CrI)** | **AP %**  **(95% CrI)** | **Rate per 100,000**  **(95% CrI)** | **Prescriptions**  **(95% CrI)** | **AP %**  **(95% CrI)** | **Rate per 100,000**  **(95% CrI)** |
| **0-5 m** | 6,880*  (3,211-10,167) | 11  (5-17) | 2,100  (984-3,100) |  |  |  | 9,184*  (5,819-12,671) | 15  (9-21) | 2,805  (1,781-3,870) |
| **6-23 m** | 65,535*  (45,035-86,144) | 10  (7-13) | 6,580  (4,522-8,651) |  |  |  | 115,315*  (80,856-150,746) | 18  (12-23) | 11,578  (8,118-15,132) |
| **2-4 y** | 72,500  (29,222-119,821) | 8  (3-13) | 3,497  (1,405-5,782) |  |  |  | 210,956« | 22 | 10,175« |
| **5-14 y #** | 55,860  (-41,759-156,160) | 4  (-3-12) | 857  (-637-2,397) | **-** | **-** | - | 89,828« | 7 | 1,378« |
| **15-44 y** | 95,554  (6,792-185,445) | 3  (0-5) | 447  (31-868) | 95,554  (6,792-185,445) | 3  (0-5) | 447  (31-868) | 26,442  (-99,224-141,995) | 1  (-3-4) | 124  (-465-665) |
| **45-64 y** | 86,608  (-16,608-191,877) | 2  (0-6) | 612  (-117-1,357) | 121,570  (27,099-214,454) | 3  (1-6) | 859  (191-1,515) | 84,469  (-27,732-193,481) | 2  (-1-6) | 597  (-200-1,367) |
| **65-74 y** | 107,893  (60,407-158,507) | 5  (3-7) | 1,972  (1,104-2,901) | 138,265  (89,746-186,255) | 6  (4-8) | 2,527  (1,635-3,410) | 134,198  (72,643-193,903) | 6  (3-9) | 2,453  (1,320-3,541) |
| **≥75 y** | 149,078  (93,733-206,045) | 6  (4-8) | 3,279  (2,050-4,532) | 175,083  (119,232-232,918) | 7  (5-9) | 3,852  (2,617-5,125) | 167,780  (96,502-235,420) | 7  (4-10) | 3,691  (2,119-5,188) |

AP =Attributable proportion, CrI = credible interval, m = months, y = years. – = model struggled to run, # = Average annual RSV-attributable prescriptions for 5-14 years were estimated from 2015 to 2017 and assumed to apply to 2018, as 2018 data was excluded for this group (see model fitting). *= Prescriptions are estimated assuming English mid-year populations are equally distributed by month of age as ONS population estimates are only provided by year of age, « = Only average simulations are provided by the predict function as 1000 simulations couldn’t be produced using PostSim.
